# GLP-1 Receptor Agonists vs SGLT2 Inhibitors for Alzheimer’s Disease Risk in Type 2 Diabetes: A Systematic Review and Meta-Analysis

**DOI:** 10.64898/2026.07.22.26358702

**Authors:** Levi Leal Silva, Maria Eduarda Scheffer Valverde, Anderson Matheus Pereira Silva, Luana Diniz Guerra Braz, Kauã Kunigami da Silva, Emanuel Medeiros Aguiar, Vitor Drummond Volta, Clara de Oliveira, Renê Rodrigues Alves, Filipi Fim Andreão, Carolina Braga Moura, Diogo Haddad Santos

## Abstract

**Purpose:** People with type 2 diabetes mellitus (DM2) have an increased risk of Alzheimer’s Disease (AD). GLP-1 receptor agonists (GLP-1RAs) and SGLT2 inhibitors (SGLT2i) have emerged as potential interventions to prevent AD and dementia and may provide insights into disease mechanisms.

**Methods:** PubMed, Embase, and Cochrane were searched for randomized controlled trials (RCTs), cohort, and case-control studies between January 1, 2006, and April 15, 2025, comparing GLP-1RAs and/or SGLT2i in adults with DM2 without cognitive impairment at baseline. A total of 12 studies met the inclusion criteria (1 RCT and 11 cohort studies). The primary outcome was incident AD and dementia, assessed primarily through adjusted hazard ratio (HR). Secondary outcomes were HbA1C levels and their relation to cognitive protection. A regression analysis was performed to assess the variables that could explain the result obtained.

**Results:** GLP-1RAs and SGLT2i consistently demonstrated a reduced risk of AD (HR = 0.72; 95% CI, 0.59–0.87) and dementia (HR = 0.84; 95% CI, 0.76–0.92) compared to other glucose-lowering therapies, with moderate/high heterogeneity and moderate certainty. Also, SGLT2i showed a modest benefit over GLP-1RAs in protecting against AD (HR = 1.07; 95% CI, 1.00–1.14; p = 0.0496), and baseline HbA1c was associated with AD risk estimates among SGLT2i users but not among GLP-1RA users.

**Conclusion:** These findings reinforce the potential of GLP-1RAs and SGLT2i as therapeutic options to slow cognitive decline in DM2, possibly depending on glycemic control and central modulation.

## 1. Background

Alzheimer’s Disease (AD) is the most common type of dementia, characterized by brain atrophy, intracellular deposition of misfolded tau protein in neurofibrillary tangles, and the accumulation of extracellular amyloid-β (Aβ) protein in senile plaques [1–2]. Recent studies have shown an increased risk of AD in patients with type 2 diabetes mellitus (DM2), with evidence that hyperglycemia and insulin resistance are related to hippocampal atrophy and amyloid accumulation, supporting its role in classical AD pathophysiology [3–5]. Given the social and economic burdens associated with diabetes and its increasing global prevalence, assessing the impact of antidiabetic therapies as possible disease-modifying therapies on reducing dementia prevalence is crucial.

Currently, *glucagon-like peptide-1 receptor agonists* (GLP-1RAs) and *sodium-glucose co-transporter 2 inhibitors* (SGLT2i) have emerged in clinical guidelines as relevant antidiabetic interventions with a preventive effect on mild cognitive impairment and AD [6–7]. Although both classes show neuroprotective potential, their comparative effectiveness and Central Nervous System (CNS) mechanisms remain unclear. Beyond glycemic control, experimental and clinical evidence suggest that both drug classes may attenuate neuroinflammation, oxidative stress, and other pathways implicated in diabetes-related neurodegeneration, supporting their investigation as potential neuroprotective therapies [8].

Thus, we performed a systematic review and meta-analysis of the comparative central effects of GLP-1RAs and SGLT2i in DM2 patients with no cognitive impairment at baseline to assess the risk of AD and dementia. Our secondary aim was to explore the potential mediation by glycemic control and class-specific adverse effects.

## 2. Methods

This study adhered to the Preferred Reporting Items for Systematic Reviews and Meta-analyses (PRISMA) statement; ethical approval was not required. The PRISMA checklist is provided in **Appendix 1**. This review was registered on the International Prospective Register of Systematic Reviews (PROSPERO) under registration no. CRD420251054597 and no deviations from the protocol were noted.

### 2.1. Search strategy

We systematically searched PubMed, Embase, and Cochrane for studies published using the following medical subject heading (MeSH) terms: *“Diabetes Mellitus”, “type 2 diabetes”, “type II diabetes”, “diabetes mellitus type 2”, “T2DM”, “DM2”, “DM II”, “diabetes”, “SGLT2 inhibitors”, “SGLT2i”, “sodium-glucose co-transporter 2 inhibitors”, “empagliflozin”, “dapagliflozin”, “canagliflozin”, “ertugliflozin”, “tofogliflozin”, “ipragliflozin”, “luseogliflozin”, “bexagliflozin”, “GLP-1 receptor agonists”, “GLP-1RA”, “glucagon-like peptide-1 receptor agonist”, “liraglutide”, “semaglutide”, “dulaglutide”, “exenatide”, “lixisenatide”, “albiglutide”, “efpeglenatide”, “Alzheimer’s disease”, “Alzheimer”, “AD”, “dementia”, “cognitive decline”, “cognitive impairment”, “neurodegenerative disease”, “mild cognitive impairment”, “Alzheimer’s disease-related dementias”, and “ADRD”.* The full search strategy employed in each database is outlined in **Supplementary Table 1**.

### 2.2. Eligibility criteria

#### 2.2.1 Inclusion criteria

We included studies that met the following eligibility criteria: (1) adult and elderly patients with DM2, (2) GLP-1 receptor agonists and/or SGLT2i compared to other antidiabetic drugs, placebo, or no treatment, (3) without cognitive impairment or a diagnosis of AD or dementia at the baseline, (4) had a follow-up duration of 24 weeks or more, and (5) comprised cohort studies, case-control studies, and randomized controlled trials (RCTs) with any number of participants. These study designs were preferred in order to provide a better understanding of correlation during the longitudinal evaluation, which was measured by adjusted hazard ratios (aHR). Two independent reviewers (V.V. and L.L.) screened the search results by title, abstract, and full text, with discrepancies discussed and resolved between the authors when necessary. The references of included studies and systematic reviews were examined for additional relevant studies.

#### 2.2.2 Exclusion criteria

Exclusion criteria included: (1) patients with relevant cognitive impairment at baseline, (2) non-human studies, and (3) narrative reviews, cross-sectional studies, case reports, comments, or letters to the editor without primary data. Only studies published in English after 2006 were included, with the search conducted from January 1, 2006, to April 15, 2025. Initially, the preprint version of Anagnostakis 2025 was included, but it was replaced with the full article after its 2026 publication. The focus of our analysis was on studies conducted between 2014 and 2025, as this period reflects the emergence of robust evidence regarding the cardiovascular, renal, and neurological benefits of GLP-1RAs and SGLT2i, particularly about AD risk in individuals with DM2 [9–10]. After the initial search, two authors (L.L. and V.V.) removed duplicates, screened titles and abstracts, and evaluated full-text articles for eligibility. A consensus-based discussion resolved discrepancies.

### 2.3. Data extraction

Two authors (L.L. and C.O.) independently extracted the following data from published articles baseline characteristics and outcome data using prespecified criteria regarding study design (*first author, year of publication, design, country*) and patient characteristics (*sample size, case-control distribution, gender, age, medication, comorbidities, polysubstance use, HbA1C levels, blood pressure, and body mass index*), interventions, follow-up, and outcomes (Montreal Cognitive Assessment [*MoCA] score, Hazard ratios [HR] of AD’s risk,* glycemia and glycated hemoglobin [*HbA1C] levels at follow-up, and risk factors*) for a cognitive assessment. The extracted information was subsequently reviewed by three additional investigators (L.B., K.K., and E.M.). Disagreements were resolved by consensus among the authors. Furthermore, Abdullah 2024 reported patients’ HbA1C levels at baseline in different units, with values that deviated from the diagnostic standards for diabetes, some of which were at levels well below physiological levels. We contacted the authors to clarify potential data inconsistencies, but no response was received.

### 2.4. Outcomes

The primary outcome was prespecified as a reduction of AD and dementia risk to evaluate the neurodegenerative progression of GLP-1RAs and/or SGLT2i in DM2 patients compared to other antidiabetic drugs. To standardize central effect measures, HR was defined to evaluate the incidence of AD and dementia. When available, MoCA scores and the Mini-Mental State Examination (MMSE) were extracted for descriptive analysis [11–12].

The secondary outcome was the relationship between AD risk and glycemic control, aimed at determining whether the neuroprotective effects are intrinsic to the drugs or mediated by metabolic improvements. This was assessed by comparing HbA1c levels with cognitive outcomes to evaluate whether reduced cognitive decline occurs independently of glucose-lowering effects. Moreover, adverse events related to GLP-1 receptor agonists and SGLT2 inhibitors were defined as a secondary outcome to inform clinical decision-making more effectively. The focus was on class-specific adverse effects, particularly those with potential neurological (such as dizziness, fatigue, mood changes, or cerebrovascular events) or metabolic relevance (including hypoglycemia, diabetic ketoacidosis, weight loss, and dehydration) [13–14]. This allows for a broader understanding of safety profiles and may help guide individualized treatment choices in patients at risk of cognitive impairment.

#### 2.4.1 Subgroup and Meta-regression Analyses

To investigate whether study-level characteristics influenced the association between antidiabetic treatment and dementia risk, we conducted *meta-regression analyses* [15]. Each meta-regression tested one moderator at a time to avoid overfitting, given the limited number of included studies in the review. Independent variables (moderators) were prespecified based on biological plausibility and clinical relevance. The following covariates were examined: (1) total sample size (n), (2) proportion of female participants (%), (3) baseline cognitive status (cognitively normal vs mild cognitive impairment), (4) dementia diagnostic specificity (Alzheimer’s dementia vs *all-cause dementia)* (5) type of antidiabetics comparison between groups and (6) HbA1c categories (*used as a proxy for glycemic control*). Effect modification was assessed based on the significance of the moderator coefficient (β) and the residual heterogeneity (I²).

#### 2.4.2 Diagnostic criteria of AD and dementia

The diagnostic criteria used by physicians for AD and dementia include International Classification of Diseases, 10th Revision (ICD-10); ICD-10 Clinical Modification (ICD-10-CM); Medical Dictionary for Regulatory Activities (MedDRA, version 21.1); Chronic Conditions Warehouse (CCW) chronic condition algorithms, or a validated algorithm (one hospitalization with a dementia record on database, or three physician claims for dementia in the database at least 30 days apart in 2 years, or a cholinesterase inhibitor prescription.

### 2.5. Risk of bias assessment and quality of evidence

The risk of bias of each included study was individually evaluated by two independent reviewers (L.B. and K.K.) using the Cochrane risk-of-bias (RoB) tool (RevMan version 5.4) for RCTs and ROBINS-I for cohort studies [16–17]. Funnel plots were constructed to visually assess potential small-study effects. Formal statistical testing using Egger’s regression was not performed due to the limited number of included studies, as recommended by the Cochrane Handbook (k < 10 for both outcomes) [18]. The quality of evidence for each outcome was then assessed using the Grading of Recommendations Assessment, Development and Evaluation (GRADE) approach [19]. To estimate the absolute effects, we applied the HRs associated with representative baseline risks obtained in studies that reported annual incidences (or, when necessary, converted from cumulative risks using an exponential model). Absolute risks were calculated using pooled baseline risks from control groups. The risk in the intervention group was estimated from hazard ratios using the formula *p₁ = 1 − (1 − p₀)^HR*, and absolute risk differences were expressed per 1,000 participants. Confidence intervals were derived by applying the lower and upper bounds of the hazard ratios to the same model.

### 2.6. Statistical analysis

The meta-analysis used a random-effects model with inverse-variance weighting based on the DerSimonian and Laird method, accounting for potential clinical and methodological heterogeneity across studies [20]. To evaluate heterogeneity among the included studies, the Cochrane Q test and I² statistic were used, with I² values of 25%, 50%, and 75% interpreted as low, moderate, and high heterogeneity, respectively [21]. To integrate direct comparisons among the antidiabetic drugs and assess their relative efficacy in preventing cognitive impairment, a meta-analysis was performed [22].

Forest plots were constructed to display individual and pooled effect estimates, following current methodological guidelines [23]. When applicable, prediction intervals were also calculated to estimate the range of true effects across settings. [24]. All statistical analyses were performed using R software version 4.4.2 [25]. The Cochrane Handbook for Systematic Reviews of Interventions was the methodological reference [26].

## 3. Results

### 3.1 Study selection

We identified 882 abstracts and performed full-text assessments of 33 articles (**Figure 1)**. Twelve unique studies (1,792,333 patients) met the inclusion criteria, including 1 RCT and 11 cohort studies [27–38]. Fifteen full-text articles were excluded due to the population not fitting the criteria, such as patients with cognitive impairment, such as AD or MCI, at baseline, or without DM2 at baseline. The mean age was 62.9 years, and the mean follow-up duration was 4.5 years (range: 1.5 - 8), supporting a long-term analysis. All studies were conducted exclusively in patients over 18 years old, diagnosed with DM2, who were candidates for glucose-lowering drugs. Three studies compared GLP-1RAs directly with SGLT2i (Anagnostakis 2025, Sun 2025, Tang 2025b), while two cohorts compared GLP-1RAs with glucose-lowering drugs (GLDs) other than SGLT2i (Tang 2025a, Tang 2025b). Since the Tang 2025b study presents three distinct cohorts with no overlap in patients, all were included in the analyses and classified as (A) GLP-1RAs versus second-line GLDs, (B) GLP-1RAs versus SGLT2i, and (C) SGLT2i versus second-line GLDs. Three cohorts analyze SGLT2i versus dipeptidyl peptidase-4 inhibitors (DPP4i) (Abdullah et al. 2025, Wu 2023, Zhuo 2025), while another two describe SGLT2i in a single-arm design (Kim 2024, Siao 2022). Finally, two more studies reported a GLP-1RAs single-arm (Nørgaard 2022, Zhou 2021), in addition to pooled data from three RCTs (Nørgaard 2022).

**Figure 1.**
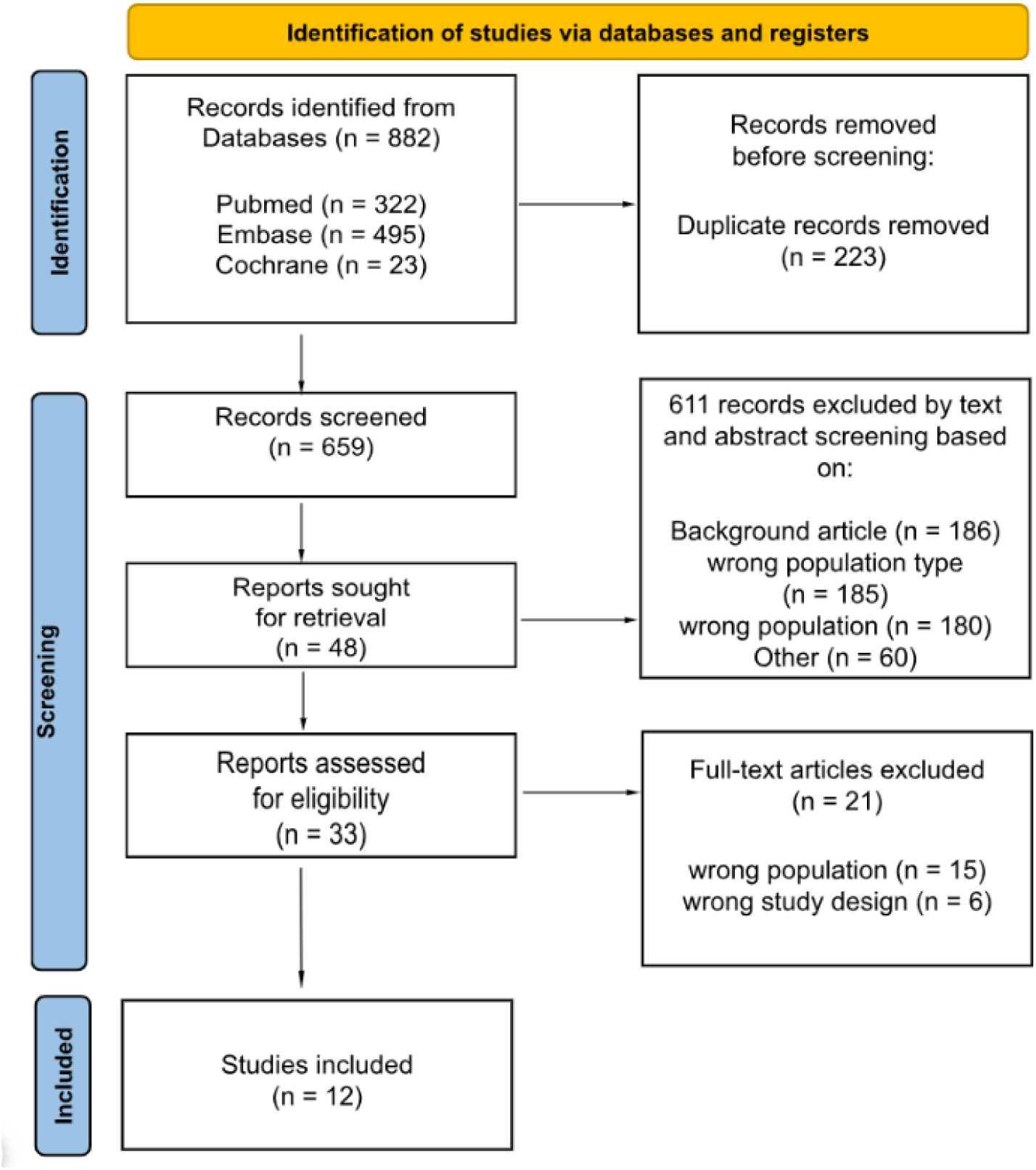
PRISMA flow diagram of study screening and selection.

The polysubstance use among the sample size includes other antidiabetic drugs, statins, diuretics, anticoagulants, angiotensin-converting enzyme inhibitors, steroids, antidepressants, and others. In addition, obesity, hypertension, dyslipidemia, and cardiovascular diseases were comorbidities significantly prevalent at the baseline in the patients included, which can be risk factors for cognitive impairment and neurodegenerative progression. Given these potential confounding factors, plots were generated from the adjusted hazard ratio, which is more precisely expressed in the Cox regression model, highlighting the central effect of GLP-1RAs and SGLT2i. A comprehensive description of the baseline patient and study characteristics is presented in **Table 1**.

**Table 1.**
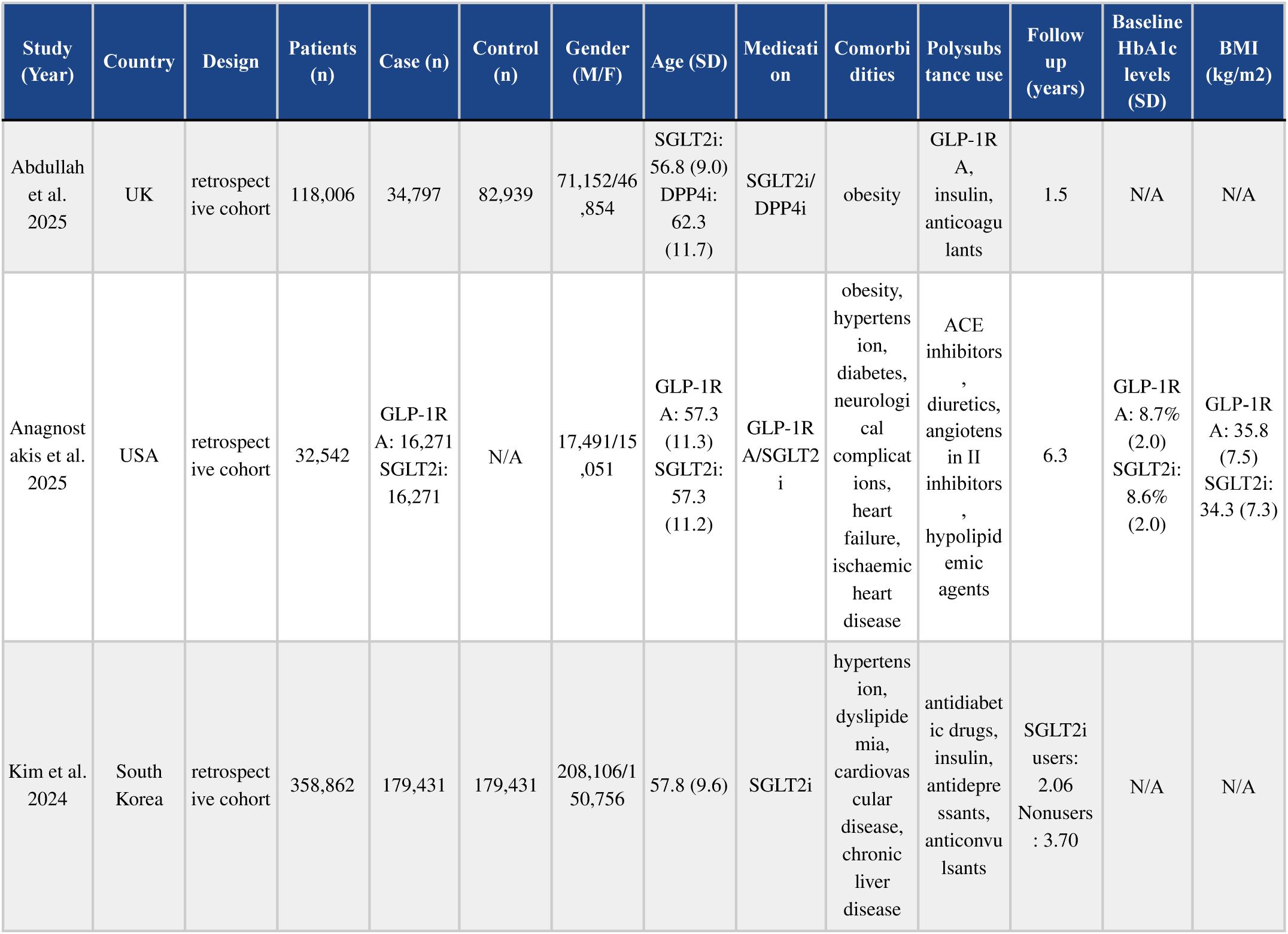
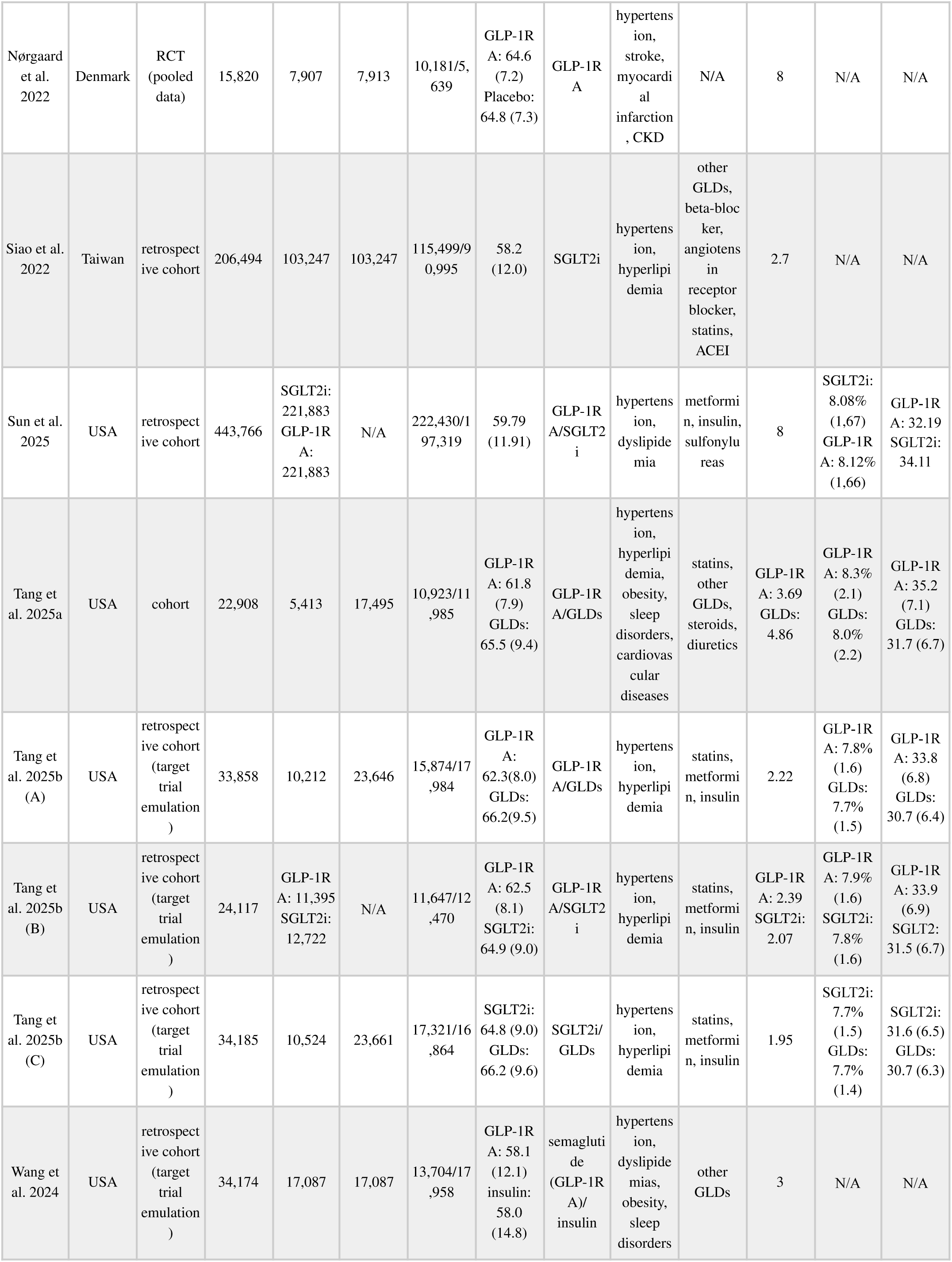
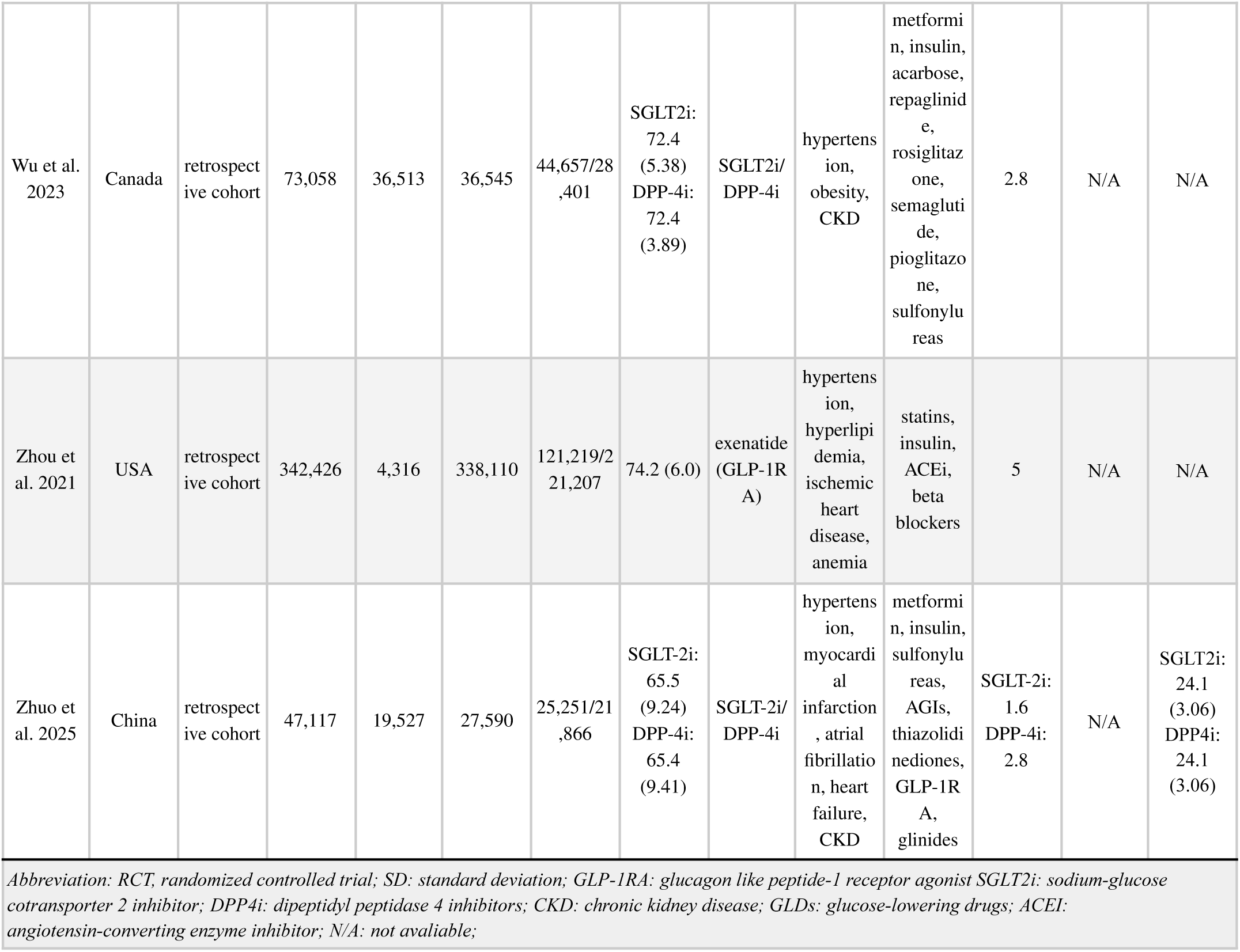
Baseline patient and study characteristics.

### 3.2 Overall risk of AD and dementia

Across twelve studies, the use of antidiabetic agents was associated with a significantly reduced risk of Alzheimer’s disease compared with controls (HR = 0.72; 95% CI, 0.59–0.87; p < 0.0001; **Figure 2**), corresponding to an approximate 28% relative risk reduction. This effect was statistically significant under a random-effects model and remained consistent despite moderate-to-high between-study heterogeneity (I² = 82.3%). Therefore, the overall risk for dementia in the same twelve studies also demonstrates a reduction of 16% in favor of the antidiabetic group (HR = 0.84; 95% CI, 0.76–0.92; p < 0.001; **Figure 2**). The random-effects model used inverse-variance weighting and yielded a statistically significant overall effect (Z = –3.81). The study outcomes of the included articles are individually listed in **Table 2**. In addition, stratified subanalyses for GLP-1RAs and SGLT2 inhibitors and their respective risks of AD and dementia are presented in **Figures S1–S5** in the supplementary material, including a direct comparison between SGLT2 inhibitors and DPP-4 inhibitors regarding the risk of dementia shown in **Figure S5**.

**Figure 2.**
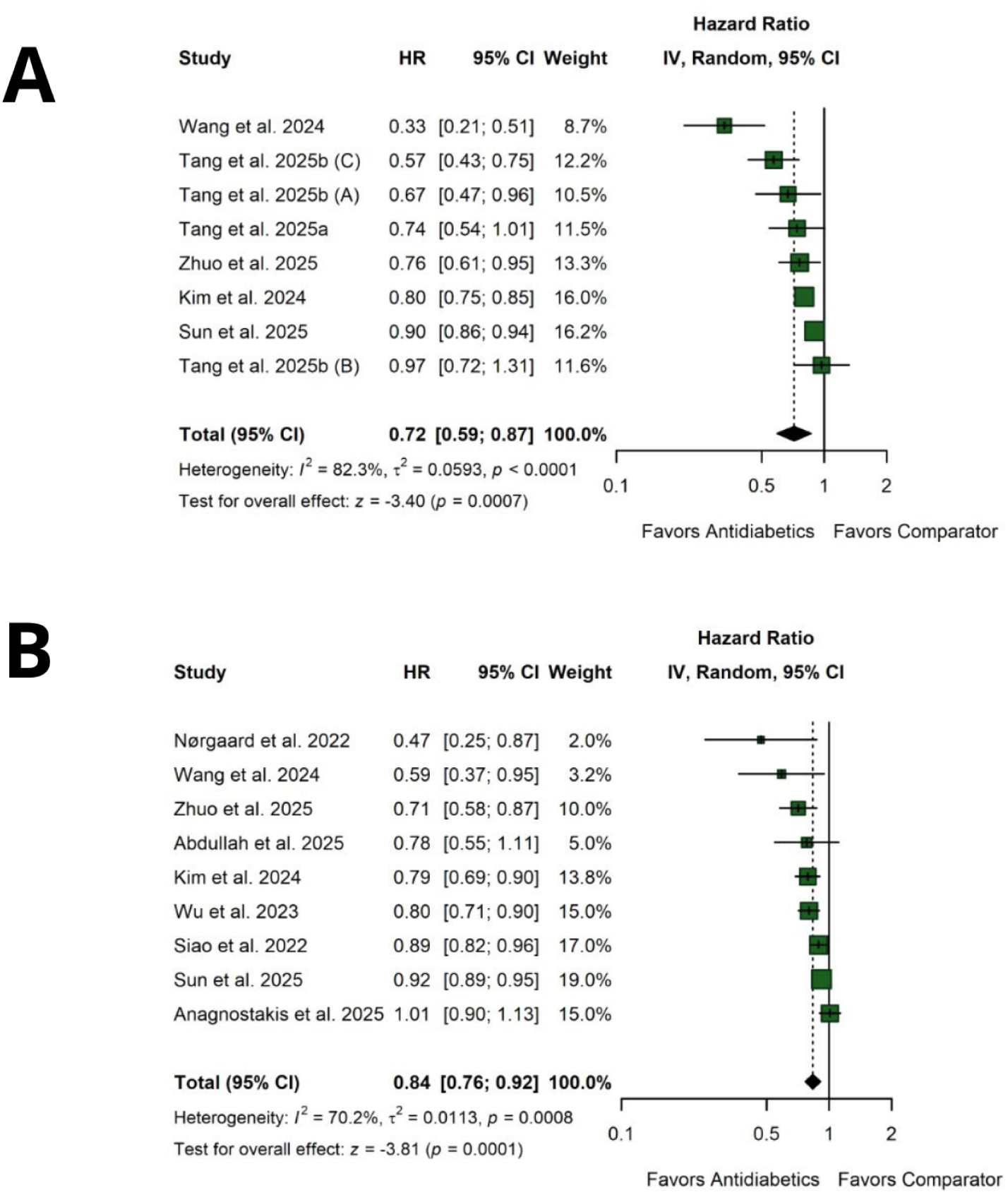
(A): Risk of AD associated with antidiabetic therapy. Forest plot of hazard ratios (HRs) with 95% confidence intervals (CIs) comparing GLP-1RAs and/or SGLT2i versus controls. (B): Risk of dementia associated with antidiabetic therapy. Forest plot of hazard ratios (HRs) with 95% confidence intervals (CIs) comparing GLP-1RAs and/or SGLT2i versus controls.

**Table 2.** Study outcomes.

| Study (Year) | Patients (n) |  | Development of AD (n) |  | Development of dementia (n) |  | Medication | Risk of AD (HR) | Risk of dementia (HR) | Risk factors |
| --- | --- | --- | --- | --- | --- | --- | --- | --- | --- | --- |
|  | Case (n) | Control (n) | Case | Control | Case | Control |  |  |  |  |
| Abdullah et al. 2025 | 34,797 | 82,939 | N/A | N/A | 40 | 533 | SGLT2i/DPP-4i | N/A | 0.78 (0.55–1.12) | comorbidities, medication use, smoking, alcohol |
| Anagnostakis et al. 2025 | GLP-1RA: 16,271<br>SGLT2i: 16,271 | N/A | N/A | N/A | GLP-1RA: 581<br>SGLT2i: 572 | N/A | GLP-1RA/SGLT2i | N/A | 1.01 (0.9–1.13) | N/A |
| Kim et al. 2024 | 179,431 | 179,431 | 1,474 | 4,244 | 1,544 | 4,454 | SGLT2i | 0.80 (0.75–0.85) | 0.79 (0.69–0.90) | N/A |
| Nørgaard et al. 2022 | 7,907 | 7,913 | N/A | N/A | 15 | 32 | GLP-1RA | N/A | 0.47 (0.25–0.86) | comorbidities |
| Siao et al. 2022 | 103,247 | 103,247 | N/A | N/A | 3,426 | 4,507 | SGLT2i | N/A | 0.89 (0.82–0.96) | N/A |
| Sun et al. 2025 | SGLT2i: 221,883<br>GLP-1RA: 221,883 | N/A | GLP-1RA: 4,244<br>SGLT2i: 3,225 | N/A | GLP-1RA: 6,063<br>SGLT2i: 7,661 | N/A | GLP-1RA/SGLT2i | 0.90 (0.86–0.94) | 0.92 (0.89–0.95) | comorbidities, medication use |
| Tang et al. 2025a | 5,413 | 17,495 | 55 | 458 | N/A | N/A | GLP-1RA/GLDs | 0.74 (0.52–0.96) | N/A | comorbidities, alcohol, smoking |
| Tang et al. 2025b (A) | 10,212 | 23,646 | 75 | 739 | N/A | N/A | GLP-1RA/GLDs | 0.67 (0.47–0.96) | N/A | comorbidities, alcohol, smoking |
| Tang et al. 2025b (B) | GLP-1RA: 11,395<br>SGLT2i: 12,722 | N/A | GLP-1RA: 90<br>SGLT2i: 130 | N/A | N/A | N/A | GLP-1RA/SGLT2i | 0.97 (0.72–1.32) | N/A | comorbidities, alcohol, smoking |
| Tang et al. 2025b (C) | 10,524 | 23,661 | 101 | 642 | N/A | N/A | SGLT2i/GLDs | 0.57 (0.43–0.75) | N/A | comorbidities, alcohol, smoking |
| Wang et al. 2024 | 17,087 | 17,087 | 27 | 73 | N/A | N/A | semaglutide (GLP-1RA)/insulin | 0.33 (0.21–0.51) | 0.59 (0.37–0.95) | alcohol, obesity, tobacco |
| Wu et al. 2023 | 36,513 | 36,545 | N/A | N/A | 560 | 696 | SGLT2i/DPP-4i | N/A | 0.80 (0.71–0.89) | comorbidities |
| Zhou et al. 2021 | 4,316 | 338,110 | 204 | 26,778 | N/A | N/A | exenatide (GLP-1RA) | N/A | N/A | comorbidities |
| Zhuo et al. | 19,527 | 27,590 | 116 | 336 | 133 | 391 | SGLT-2i/DP | 0.76 | 0.71 | comorbidities |
| 2025 |  |  |  |  |  |  | P-4i | (0.61–0.96) | (0.58–0.87) | s |

Abbreviation: GLP1RA: glucagon like peptide-1 receptor agonist SGLT2i: sodium-glucose cotransporter 2 inhibitor; HR: hazard ratio; N/A: not available;

In addition, we performed an overall HR analysis for dementia and a sensitivity analysis only in studies with low risk of bias to evaluate the data from the initial analyses with greater certainty. As reported in **Figure S6**, the overall risk for dementia in the three studies with low risk demonstrates a reduction of 18% in favor of the antidiabetic group (HR = 0.82; 95% CI: 0.71–0.95). The overall HR analysis for AD of these studies was not possible because only Kim 2024 and Nørgaard 2022 reported data.

#### 3.2.1 Exploration of effect modifiers in meta**-**analysis

Meta-regression analyses were conducted to investigate whether study-level characteristics accounted for variability in the estimated hazard ratios for AD. Six models were fitted, each testing one moderator at a time, as described in **Supplementary Table 2**. Overall, none of the evaluated moderators showed a statistically significant association with hazard ratio estimates, and most models demonstrated persistent moderate-to-high residual heterogeneity. The proportion of female participants was not associated with effect estimates (β = −0.016; 95% CI −0.044 to 0.013; p = 0.275), explaining only a small fraction of between-study variance (R² = 5.9%), with substantial residual heterogeneity (I² = 75.9%). Similarly, baseline cognitive status (β = −0.289; 95% CI −0.826 to 0.247; p = 0.29; R² = 9.8%; I² = 78.8%) and dementia classification (β = 0.119; 95% CI −0.111 to 0.349; p = 0.311; R² = 21.9%; I² = 75.2%) were not significantly associated with hazard ratios, indicating limited explanatory capacity.

Comparator type also did not significantly explain variability in effect estimates (QM = 4.70; p = 0.319), with residual heterogeneity remaining moderate (I² ≈ 49.5%) and explained only a small proportion of the variance (R² ≈ 6.3%). Sample size (log-transformed) showed a non-significant trend of association (β = 0.064; 95% CI −0.011 to 0.138; p = 0.096), accounting for a larger proportion of the heterogeneity (R² = 50.6%), although residual variability persisted (I² = 64.1%). Regarding glycemic control, baseline HbA1c levels were not significantly associated with risk ratios in the GLP-1RAs model (β = 0.439; 95% CI −0.573 to 1.451; p = 0.395), explaining a minimal proportion of heterogeneity (R² = 7.6%) with moderate residual variability (I² = 58.6%). In contrast, the SGLT2i model demonstrated a statistically significant association (β = 1.127; 95% CI 0.534 to 1.719; p < 0.001), with complete reduction of residual heterogeneity (I² = 0%; R² = 100%); however, this finding should be interpreted with caution, given the limited number of studies included in this model.

Despite exploring multiple potential moderators, several models demonstrated substantial residual heterogeneity (I² > 65%) with limited explanatory value. This suggests that the examined study-level variables, including sex distribution, cognitive status classification, diagnostic specificity, and comparator type, do not fully account for the variability in hazard ratios across studies. The persistent between-study variance likely reflects unmeasured differences in population characteristics, duration of follow-up, baseline cognitive trajectories, or methodological heterogeneity not captured by the selected moderators. Meta-regression analyses for the risk of AD were completely reported in **Supplementary Table 3**. Full model outputs are detailed in the Supplementary Material, and all corresponding meta-regression plots are provided in **Figures S7–S12**.

### 3.3 GLP-1RAs vs SGLT2i

To evaluate the differential neuroprotective effect of GLP-1RAs and SGLT2i on AD risk, a head-to-head comparison was conducted using the adjusted hazard ratios (aHRs) from studies directly comparing both classes. A random-effects model, as presented in **Figure 3**, was applied due to the limited number of studies.

**Figure 3.**
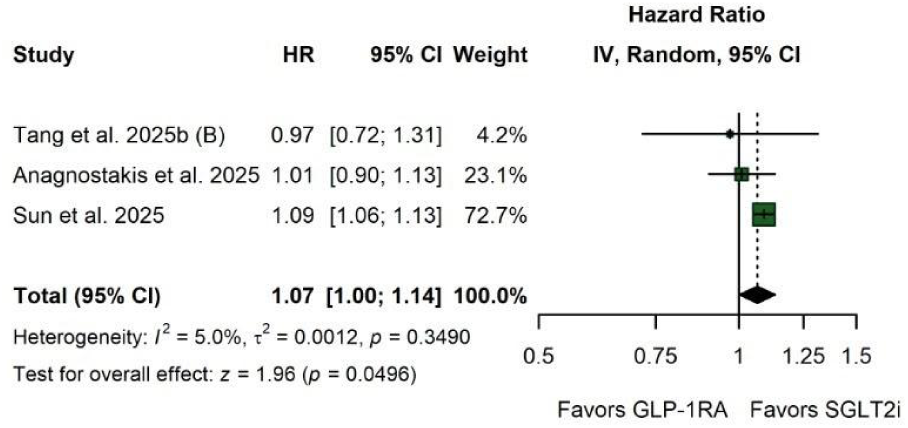
Direct comparison between GLP-1RAs vs SGLT2i. Forest plot of hazard ratios (HRs) with 95% confidence intervals (CIs) comparing GLP-1RAs versus SGLT2i, which was being standardized as the control group.

Although only Sun et al. 2025 demonstrated a statistically significant difference individually, the pooled analysis favored SGLT2i. The pooled analysis showed a statistically significant but marginal difference in AD risk reduction between SGLT2i and GLP-1RAs (HR = 1.07; 95% CI, 1.00–1.14; p = 0.0496), suggesting a slightly greater benefit from SGLT2i. Low heterogeneity was observed (I² = 5%, *t*^2^ = 0.0012; p = 0.34), with an overall effect test z = 1.96, suggesting consistent findings across studies, but caution is warranted due to the small number of studies included. Also, included studies provided limited data regarding adverse effects, precluding quantitative synthesis.

### 3.4 Sensitivity Analyses

Leave-one-out sensitivity analyses were performed to assess the robustness of the association between antidiabetic treatment and the risk of AD and dementia. Each iteration recalculated the pooled HR after omitting one study at a time. The overall effect for AD remained stable (pooled HR = 0.85; 95% CI: 0.79–0.92; I² = 70.0%), indicating that no single study exerted a disproportionate influence on the summary estimate (**Figure S13**). Variations in heterogeneity were observed across iterations (I² range: 50.5%–78.0%), but these did not meaningfully affect the direction or magnitude of the pooled effect.

A similar pattern was observed for dementia (**Figure S14**), with consistent effect estimates across leave-one-out analyses (pooled HR = 0.87; 95% CI: 0.80–0.95; I² = 77.7%). Heterogeneity varied more widely across iterations (I² range: 0.1%–73.8%), suggesting that specific studies contributed disproportionately to between-study variability; however, the overlap of confidence intervals and the relatively low between-study variance (τ² ≈ 0.012) indicate that the overall findings remain robust. Sensitivity analysis restricted to studies with low risk of bias (**Figure S15**) yielded comparable results (pooled HR = 0.82; 95% CI: 0.71–0.95; I² = 66.4%), with moderate heterogeneity across leave-one-out iterations (I² range: 56.3%–74.8%). These findings support the stability of the main results and suggest that the observed associations are not driven by studies at higher risk of bias.

### 3.5 Quality Assessment and risk of bias

The largest contributions to the model came from Sun 2025, Siao 2022, and Kim 2024, which together accounted for more than 50% of the total weight. In contrast, studies with low precision, such as Abdullah 2025, exerted minimal influence on the summary estimate. The primary forest plots are presented in **Figure 2**, while publication bias is reported on funnel plots in **Figures S16-S17**. The Risk of Bias tool version 2 (RoB 2) and the Risk of Bias In Non-randomized Studies of Interventions (ROBINS-I) were used to assess the risk of bias in the pooled RCT data and the 11 cohort studies, respectively.

As shown in **Supplementary Table 4**, we summarized the main outcomes and assessed the certainty of evidence using the GRADE approach. Although hazard ratios consistently suggested relative risk reductions, the corresponding absolute risk differences were modest, ranging from approximately 3 to 9 fewer cases of dementia or AD per 1,000 person-years. Accordingly, outcomes A and B were rated as moderate certainty of evidence, primarily due to downgrading by one level for risk of bias, reflecting the inclusion of observational studies with moderate risk of bias. No further downgrading was applied for inconsistency, indirectness, imprecision, or publication bias. In contrast, outcome C (GLP-1RAs versus SGLT2i) was rated as low certainty of evidence due to downgrading for both risk of bias and imprecision, the latter reflecting the small number of included studies and limited precision of the estimates. These factors may lead to an overestimation of the comparative effect between GLP-1RAs and SGLT2 inhibitors. No upgrading criteria were met, as the included studies did not demonstrate a large magnitude of effect, evidence of a dose-response relationship, or residual confounding likely to reduce the observed effect. Overall, the consistency of adjusted analyses across studies reduces the likelihood that unmeasured confounding alone explains the observed associations, although this possibility cannot be entirely excluded.

Due to the lack of adjustment for all relevant confounders, specific analyses to estimate the effect of assignment to intervention, and prior publication of protocols with prespecified outcomes, D1, D4, and D7, respectively, showed significant bias (**Figures S18-S19**). Specifically, D2, D3, and D5 presented a risk of bias in the assignment of interventions to groups, in the selection of eligible patients for the cohort, and in the availability of essential participant data, respectively. Each domain was classified as having low, moderate, serious, or critical risk of bias, depending on the severity that the lack of information or gap in the study design could imply. Otherwise, the three RCTs pooled in Nørgaard 2022 were individually assessed and identified by the RoB 2 tool as being of low risk of bias.

## 4. Discussion

In this systematic review and meta-analysis, GLP-1RAs and SGLT2 inhibitors were associated with a 28% reduction in the risk of AD and a 16% reduction in the risk of dementia at any point during follow-up, with a marginal benefit of SGLT2 inhibitors over GLP-1 receptor agonists in reducing the risk of AD. Exploratory meta-regression revealed no significant association between baseline HbA1c and hazard ratios for AD among GLP-1RAs users, whereas a statistically significant positive association was observed for SGLT2i, suggesting that the mechanisms underlying neuroprotection may differ between classes. To our knowledge, this is the first review to conduct a head-to-head comparison of GLP-1RAs versus SGLT2i. A statistically significant 7% difference was observed between SGLT2i and GLP-1RAs for AD prevention, in contrast to data from a recent RCT [39]. Together, these findings support a neuroprotective role for both agents in DM2 patients, while raising important questions about the pathways through which this benefit is conferred.

Compared with non-users or users of other glucose-lowering drugs, GLP-1RAs and SGLT2i users demonstrated a consistent reduction in AD and dementia risk across included studies. A previous network meta-analysis reported odds ratios of 0.41 and 0.34 for dementia risk with SGLT2i and GLP-1RA, respectively — substantially lower than the hazard ratio of 0.84 observed in our analysis [40]. This discrepancy likely reflects differences in study design, follow-up duration, and the metrics used, but also raises the possibility that the neuroprotective effect of these agents accumulates over time, exerting a progressively greater benefit on synaptic integrity as metabolic dysregulation is sustained or corrected. The chronic sequelae of DM2 — including oxidative stress, microangiopathy, neuroinflammation, and diabetic encephalopathy — represent a slowly evolving biological substrate, and their prevention may require long-term metabolic regulation to translate into measurable cognitive protection [41–42].

The primary outcomes of our analysis span the full cognitive impairment spectrum, from MCI to dementia, encompassing the neurodegenerative cascade of AD characterized by pathological accumulation of β-amyloid and tau protein aggregates, typically accompanied by a prominent amnestic pattern of cognitive decline [40, 43–44]. Notably, a substantial proportion of patients with MCI progress to dementia not solely due to irreversible neurodegeneration, but partly driven by the persistence of modifiable risk factors. Those who fail to implement lifestyle changes — such as improved diet, physical activity, and glycemic control — are more likely to develop or worsen DM2, thereby increasing their dementia risk through mechanisms including oxidative stress, microangiopathy, chronic neuroinflammation, and diabetic encephalopathy. Conversely, patients who adhere to lifestyle modifications achieve better metabolic control and may attenuate this progression. In this context, patients benefiting from GLP-1RAs in terms of dementia risk reduction may already be experiencing significant weight loss — raising the question of whether the observed neuroprotective effects are mediated directly by the drug or are secondary to systemic metabolic improvement.

Whether antidiabetic agents exert a direct effect on neuroinflammation or whether their neuroprotective benefits are entirely secondary to glycemic reduction remains an open question — and it is plausible that both mechanisms operate simultaneously [45–46]. SGLT2 inhibitors offer a particularly informative perspective in this regard: unlike GLP-1RA, they do not significantly promote weight loss, yet still achieve meaningful glycemic control and have demonstrated comparable associations with reduced dementia risk. This observation suggests that glycemic regulation itself may represent a central mechanistic pathway, independent of weight reduction [47]. Consistent with this, pre-clinical and clinical studies suggest that mechanistic pathways such as neuroinflammation and mitochondrial function may contribute independently of HbA1c and BMI [48–49]. In particular, semaglutide has been associated with a reduction in neuroinflammation severity through central neuronal GLP-1 receptors [50], and GLP-1RAs may cross the blood-brain barrier to directly modulate tau phosphorylation and synaptic plasticity [51–53] — hallmarks of AD progression. In a study of cultured rat hippocampal neurons, Aβ oligomers transiently inhibited AMPK translocation to the cell membrane, reducing GLUT4-mediated glucose uptake; insulin prevented this inhibition, suggesting that antidiabetic drugs may exert significant neuroprotective effects through metabolic modulation [54]. These findings collectively support a dual mechanism: direct central action and indirect systemic metabolic benefit.

The failure of recent clinical trials of GLP-1RAs to demonstrate a reduction in clinical progression among patients with established AD does not necessarily contradict our findings [55]. Rather, it likely reflects a fundamental distinction between disease prevention and disease modification: once overt neurodegeneration is present — with advanced amyloid burden and tau pathology — pharmacological intervention may be insufficient to reverse an already established cascade [56]. The population examined in our meta-analysis consists predominantly of DM2 patients without established neurodegeneration, but with chronic metabolic dysregulation that promotes neuroinflammation — a potentially more actionable therapeutic window. This distinction underscores the importance of timing: the neuroprotective potential of GLP-1RAs and SGLT2i may be greatest when initiated early, before irreversible neurodegenerative changes occur, reinforcing the value of early metabolic intervention as a preventive strategy for dementia.

Given their neuroprotective potential and favorable metabolic profile, GLP-1RAs and SGLT2i may be considered preferred therapeutic options in DM2 patients at risk of cognitive decline. Their clinical indication will depend on the individual patient profile — particularly in terms of cardiovascular and renal function, BMI, and risk of adverse effects. Both classes reduce body weight, with GLP-1RAs demonstrating greater effects through incretin and anorexigenic mechanisms. Despite their established side-effect profiles — including gastroesophageal reflux disease and cholelithiasis for GLP-1RA, and urinary or genital tract infections, hypoglycemia, bone fracture, and acute renal impairment for SGLT2i [57–58] — their prevalence specifically in AD and dementia populations remains inconclusive due to limited data. Nonetheless, external clinical data support a favorable neurological and metabolic safety profile for both classes.

### Limitations

This study has several limitations. According to the GRADE system, the evidence supporting the reduction in AD and dementia risk with GLP-1RAs and SGLT2i was rated as moderate certainty, providing reasonable confidence in the estimates. In contrast, the direct head-to-head comparison between the two classes was rated as low certainty, mainly due to methodological limitations and the small number of comparative studies. The exploratory nature of the meta-regression analyses further limits causal inference. Additionally, the included studies varied in follow-up duration, comparator choice, and outcome definitions, contributing to residual heterogeneity. Further long-term randomized controlled trials are needed to validate these findings, better characterize the metabolic pathophysiology of AD, and investigate the relative contributions of direct central mechanisms versus systemic metabolic effects in the neuroprotective action of glucose-lowering drugs.

### Conclusion

In this systematic review and meta-analysis, GLP-1RAs and SGLT2i were associated with lower risks of AD and dementia in individuals with DM2, with a small superiority of SGLT2i. Importantly, HbA1c levels emerged as a possible moderator of effect estimates, suggesting that glycemic control may reduce cognitive impairment. These findings support further investigation of glucose-lowering therapies as potential strategies for dementia prevention, although evidence from long-term randomized trials remains needed.

## Declarations

### Ethics Approval

Ethical approval was not required because this study was based exclusively on previously published data.

### Consent for Publication

Not applicable.

### Data availability

The datasets extracted from the included studies and the R scripts used for all analyses are available in the Supplementary Materials. Additional resources are available upon reasonable request to the corresponding author.

### Author Contributions

L.S., M.V., and E.A. wrote the manuscript. A.S., K.S., and L.B. performed the data analysis and risk of bias. C.O, V.V., and R.A. gathered the data. L.S. is the corresponding author and designed the study with C.M. and D.S., the seniors. All authors reviewed the manuscript.

## Acknowledgements

Federal University of Rio de Janeiro, Federal University of Pernambuco, University of São Paulo, and the Faculty of Medical Sciences and Health.

## Abbreviations

GLP-1RA: Glucagon-like peptide-1 receptor agonist
SGLT2i: Sodium-glucose co-transporter-2 inhibitors
DM2: Diabetes Mellitus type 2
AD: Alzheimer’s Disease
ADRD: Alzheimer’s Disease Related Dementia
DPP4i: Dipeptidyl Peptidase-4 inhibitors
GLDs: Glucose-lowering drugs
CNS: Central Nervous System
RoB-2: Risk of Bias tool version 2
ROBINS-I: Risk Of Bias In Non-randomized Studies of Interventions
SD: Standard deviation
RCT: Randomized controlled trial

