## Supplementary Material for "GLP-1 Receptor Agonists vs SGLT2 Inhibitors for Alzheimer’s Disease Risk in Type 2 Diabetes: A Systematic Review and Meta-Analysis"

***Journal submission:***

**Running title:** GLP-1RA and SGLT2i in Alzheimer's Risk
**Manuscript type:** Systematic Review and Meta-analysis
**PROSPERO ID:** CRD420251054597

**Corresponding author:** Levi Leal Silva
Department of Medicine, Federal University of Rio de Janeiro, Rio de Janeiro, Brazil

**Funding:** No funding was received to conduct this study.
**Conflicts of interest:** The authors have no relevant financial or non-financial conflicts of interest to disclose.

**1. Supplementary Tables and Figures**

**1.1** Supplementary Table 1. Search strategy across databases (PubMed, Embase, and Cochrane).

**1.2** Supplementary Table 2. Stratification of pre-coded variables for the meta-regression.

**1.3** Supplementary Table 3. Study-level characteristics and results of meta-regression analysis for hazard ratios of dementia.

**1.4** Supplementary Table 4. Summary of Findings table showing the main outcomes of the meta-analysis and the quality of evidence according to GRADE.

**1.5** Figures S1-S5: Stratified risks of AD and dementia among users of GLP-1RAs or SGLT2i.

**1.6** Figure S6: Risk of dementia associated with antidiabetic therapy in low-risk-of-bias studies.

**1.7** Figures S7–S12. Funnel plot and meta-regression bubble plots evaluating moderators of hazard ratio estimates for AD across studies.

**1.8** Figures S13-S15. Leave-one-out sensitivity analyses for each forest plot, showing the influence of individual studies on the respective pooled effect estimates.

**1.9** Figures S16-S17: Funnel plot evaluating publication bias regarding primary outcomes.

**1.10** Figures S18-19: Risk of Bias Assessment (ROBINS-I and RoB2) of all included studies.

| |
|---|
| **PubMed** |
| ("Diabetes Mellitus"[Title/Abstract] OR "type 2 diabetes"[Title/Abstract] OR "type II diabetes"[Title/Abstract] OR "diabetes mellitus type 2"[Title/Abstract] OR "T2DM"[Title/Abstract] OR "DM2"[Title/Abstract] OR "DM II"[Title/Abstract] OR diabetes[Title/Abstract]) AND ("SGLT2 inhibitors"[Title/Abstract] OR SGLT2i[Title/Abstract] OR "sodium-glucose co-transporter 2 inhibitors"[Title/Abstract] OR empagliflozin[Title/Abstract] OR dapagliflozin[Title/Abstract] OR canagliflozin[Title/Abstract] OR ertugliflozin[Title/Abstract] OR tofogliflozin[Title/Abstract] OR ipragliflozin[Title/Abstract] OR luseogliflozin[Title/Abstract] OR bexagliflozin[Title/Abstract] OR "GLP-1 receptor agonists"[Title/Abstract] OR "GLP-1RA"[Title/Abstract] OR GLP1RA[Title/Abstract] OR "glucagon-like peptide-1 receptor agonist"[Title/Abstract] OR liraglutide[Title/Abstract] OR semaglutide[Title/Abstract] OR dulaglutide[Title/Abstract] OR exenatide[Title/Abstract] OR lixisenatide[Title/Abstract] OR albiglutide[Title/Abstract] OR efpeglenatide[Title/Abstract]) AND ("Alzheimer's disease"[Title/Abstract] OR Alzheimer[Title/Abstract] OR AD[Title/Abstract] OR "dementia"[Title/Abstract] OR "cognitive decline"[Title/Abstract] OR "neurodegenerative disease"[Title/Abstract] OR "mild cognitive impairment"[Title/Abstract] OR MCI[Title/Abstract]) |
| **Embase** |

| |
|---|
| ('diabetes mellitus type 2':ti,ab OR 'type 2 diabetes':ti,ab OR 'type II diabetes':ti,ab OR 'T2DM':ti,ab OR 'DM2':ti,ab OR 'DM II':ti,ab OR diabetes:ti,ab) AND ('SGLT2 inhibitor':ti,ab OR SGLT2i:ti,ab OR 'sodium glucose co-transporter 2 inhibitor':ti,ab OR empagliflozin:ti,ab OR dapagliflozin:ti,ab OR canagliflozin:ti,ab OR ertugliflozin:ti,ab OR tofogliflozin:ti,ab OR ipragliflozin:ti,ab OR luseogliflozin:ti,ab OR bexagliflozin:ti,ab OR 'GLP-1 receptor agonist':ti,ab OR 'GLP-1RA':ti,ab OR GLP1RA:ti,ab OR 'glucagon like peptide 1 receptor agonist':ti,ab OR liraglutide:ti,ab OR semaglutide:ti,ab OR dulaglutide:ti,ab OR exenatide:ti,ab OR lixisenatide:ti,ab OR albiglutide:ti,ab OR efpeglenatide:ti,ab) AND ('Alzheimer disease':ti,ab OR Alzheimer:ti,ab OR AD:ti,ab OR dementia:ti,ab OR 'cognitive decline':ti,ab OR 'cognitive impairment':ti,ab OR 'neurodegenerative disease':ti,ab OR 'mild cognitive impairment':ti,ab OR MCI:ti,ab OR 'Alzheimer disease related dementia':ti,ab OR ADRD:ti,ab) |
| **Cochrane** |
| (diabetes mellitus type 2 OR type 2 diabetes OR T2DM OR DM2) AND ("GLP-1 receptor agonist" OR GLP-1RA OR liraglutide OR semaglutide OR dulaglutide OR exenatide OR lixisenatide OR albiglutide OR efpeglenatide) AND ("SGLT2 inhibitor" OR SGLT2i OR empagliflozin OR dapagliflozin OR canagliflozin OR ertugliflozin OR tofogliflozin OR ipragliflozin OR luseogliflozin OR bexagliflozin) AND ("Alzheimer's disease" OR Alzheimer OR AD OR dementia OR "mild cognitive impairment" OR MCI OR "cognitive impairment" OR "cognitive decline" OR "Alzheimer's disease-related dementias" OR ADRD) |

**Supplementary Table 1:** Search strategy across PubMed, Embase, and Cochrane databases, including complete Boolean logic and key terms used.

| Study (Year) | Sample Size (n) | Risk of AD (HR) | Sex (% Female) | Baseline Cognitive Status | Dementia Classification | Comparator Type | Baseline HbA1c (SD) |
|---|---|---|---|---|---|---|---|
| Kim et al. 2024 | 358,862 | 0.80 (0.75–0.85) | 42.0% | 1 | 2 | 5 | N/A |
| Sun et al. 2025 | 443,766 | 0.90 (0.86–0.94) | 44.4% | 1 | 2 | 1 | SGLT2i: 8.08% GLP1: 8.12% |
| Tang et al. 2025a | 22,908 | 0.74 (0.52 - 0.96) | 52.3% | 1 | 1 | 4 | GLP-1RA: 8.3% GLDs: 8.0% |
| Tang et al. 2025b (A) | 33,858 | 0.67 (0.47-0.96) | 53.1% | 1 | 1 | 4 | GLP-1RA: 7.8% (1.6) 7.7% (1.5) |

| | | | | | | | |
|---|---|---|---|---|---|---|---|
| Tang et al. 2025b (B) | 24,117 | 0.97 (0.72-1.32 ) | 51.7% | 1 | 1 | 1 | GLP1: 7.9% SGLT2i: 7.8% |
| Tang et al. 2025b (C) | 34,185 | 0.57 (0.43-0.75 ) | 49,3% | 1 | 1 | 5 | SGLT2i: 7.7% (1.5) GLDs: 7.7% (1.4) |
| Wang et al. 2024 | 34,174 | 0.59 (0.37 - 0.95) | 52.50% | 2 | 2 | 3 | N/A |
| Zhuo et al. 2025 | 47,117 | 0.76 (0.61–0.96 ) | 46.4% | 1 | 2 | 2 | N/A |
| Abbreviations: AD, Alzheimer's disease; HR, Hazard Ratio; HbA1c, Hemoglobin A1C; SD, Standard Deviation; ICD, International Classification of Diseases; N/A, not available. | | | | | | | |

**Supplementary Table 2:** Stratification of pre-coded variables for the meta-regression. (A): sex proportion (percentage of females) was collected in continuous values; (B): baseline cognitive status was classified as: no dementia at baseline (1) or mild cognitive impairment or unspecified cognitive status (2); (C) classification of the diagnosis of dementia was defined according to clear clinical criteria specified by ICD-10 or subsequent editions and was classified as follows: Alzheimer's Dementia, with AD assessed as a primary or secondary outcome (1); or all-cause dementia, without specification of subtype, including any form of dementia (2); (D): comparison of antidiabetic drugs between the case and control groups was classified as GLP-1RAs versus SGLT2i (1), SGLT2i versus DPP-4i (2); semaglutide versus insulin (3), GLP-1RAs versus GLDs (4), or SGLT2i versus GLDs (5); (E): baseline HbA1c levels were collected in continuous values.

| Moderator | β (SE) | 95% CI | p-value | $I^2$ (%) | $\tau^2$ | $R^2$ (%) | p (moderator) |
|---|---|---|---|---|---|---|---|
| Sex (% female) | -0.016 (0.014) | -0.044 to 0.013 | 0.275 | 75.9 | 0.014 | 5.9 | 0.275 |
| Baseline cognitive status | -0.289 (0.274) | -0.826 to 0.247 | 0.29 | 78.8 | 0.0134 | 9.8 | 0.29 |
| Dementia classification | 0.119 (0.117) | -0.111 to 0.349 | 0.311 | 75.2 | 0.0116 | 21.9 | 0.311 |
| Comparator type | — | — | — | 49.5 | 0.0139 | 6.3 | 0.319 |
| Sample size (log) | 0.064 (0.038) | -0.011 to 0.138 | 0.096 | 64.1 | 0.0073 | 50.6 | 0.096 |
| HbA1c (GLP-1RA) | 0.439 (0.517) | -0.573 to 1.451 | 0.395 | 58.6 | 0.0162 | 7.6 | 0.395 |
| HbA1c (SGLT2i) | 1.127 (0.302) | 0.534 to 1.719 | <0.001 | 0 | 0 | 100 | <0.001 |

| Abbreviations: SE: Standard Error; CI: Confidence Interval; |
|---|

**Supplementary Table 3:** Study-level characteristics and results of mixed-effects meta-regression analyses for risk of Alzheimer's Disease [AD]. Mixed-effects meta-regression models were fitted using inverse-variance weighting and restricted maximum likelihood (REML) estimation. Study-level moderators included sex distribution (female proportion), baseline cognitive status, dementia classification, comparator type, sample size (log-transformed), and baseline HbA1c levels. Effect estimates are presented as regression coefficients (β) with standard errors (SEs), 95% confidence intervals (CIs), and corresponding p-values. Between-study heterogeneity is reported using $I^2$ and $\tau^2$, and the proportion of heterogeneity explained by each model is expressed as $R^2$. P-values for moderators correspond to the omnibus test of moderators (QM). Residual heterogeneity remained moderate to high across most models.

| **Certainty assessment** | | | | | | | **No. of patients** | | **Effect** | | **Certainty** | **Importance** |
|---|---|---|---|---|---|---|---|---|---|---|---|---|
| **No. of studies** | **Study design** | **Risk of bias** | **Inconsistency** | **Indirectness** | **Imprecision** | **Publication Bias** | **GLP-1 RA/SGLT2i** | **Control** | **Relative (95% CI)** | **Absolute (95% CI)** | | |
| ***A. AD risk*** | | | | | | | | | | | | |
| 8 | Randomized trials and cohorts | Downgraded by One Level | Do not downgrade | Do not downgrade | Do not downgrade | Do not downgrade | 710,077 | 288,910 | HR 0.72 (0.59–0.87) | 6 fewer per 1,000 (3-9 fewer) | ⊕⊕⊕ **Moderate** | CRITICAL |
| ***B. Dementia risk*** | | | | | | | | | | | | |
| 9 | Randomized trials and cohorts | Downgraded by One Level | Do not downgrade | Do not downgrade | Do not downgrade | Do not downgrade | 874,817 | 454,752 | HR 0.84 (0.76-0.92) | 5 fewer per 1,000 (3 -7 fewer) | ⊕⊕⊕ **Moderate** | CRITICAL |
| ***C. GLP-1RA versus SGLT2i*** | | | | | | | | | | | | |
| 3 | Randomized trials and cohorts | Downgraded by One Level | Do not downgrade | Do not downgrade | Do not downgrade | Downgraded by One Level | 585,889 | N/A | HR 1.07 (1.00–1.14) | 2 more per 1,000 (0-4 more) | ⊕⊕ **Low** | CRITICAL |

**Supplementary Table 4:** Summary of Findings table showing the main outcomes of the meta-analysis and the quality of evidence according to GRADE. Outcome A (AD risk): Moderate certainty due to moderate risk of bias; Outcome B (Dementia risk): Moderate certainty due to moderate risk of bias; Outcome C (GLP-1RA versus SGLT2i): low risk due to moderate risk of bias and small sample of studies analyzed; Abbreviations: N/A: not available; CI: confidence interval.

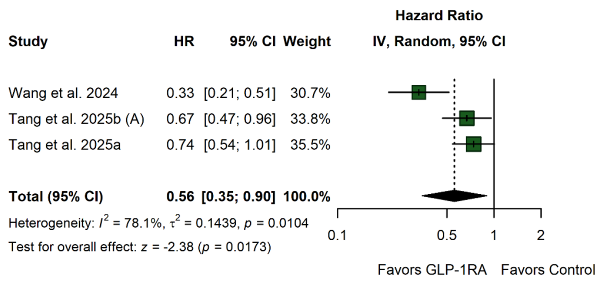


**Figure S1.** Risk of AD in GLP-1RA users. Forest plot of hazard ratios (HRs) with 95% confidence intervals (CIs) comparing GLP-1RA users versus controls across three studies. Pooled estimates were obtained using a random-effects model with inverse-variance weighting. Squares represent individual study effects (scaled by weight), horizontal lines indicate 95% CIs, and the diamond represents the pooled effect. Values <1 favor GLP-1RA. Substantial heterogeneity was observed ($I^2 = 78.1\%$; $\tau^2 = 0.1439$; $p = 0.0104$).

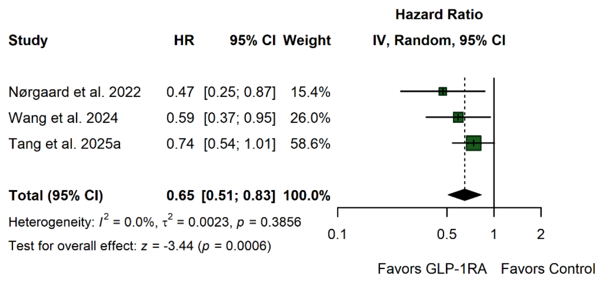


**Figure S2.** Risk of dementia in GLP-1RA users. Forest plot of hazard ratios (HRs) with 95% confidence intervals (CIs) comparing GLP-1RA users versus controls across three studies. Pooled estimates were obtained using a random-effects model with inverse-variance weighting. Squares represent individual study effects (scaled by weight), horizontal lines indicate 95% CIs, and the diamond represents the pooled effect. Values <1 favor GLP-1RA. No substantial heterogeneity was observed ($I^2 = 0.0\%$; $\tau^2 = 0.0023$; $p = 0.3856$).

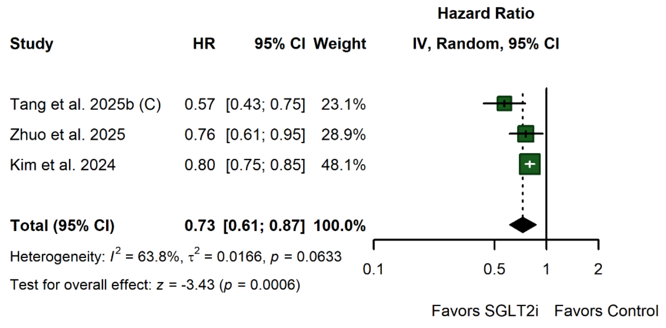


**Figure S3.** Risk of AD in SGLT2i users. Forest plot of hazard ratios (HRs) with 95% confidence intervals (CIs) comparing SGLT2i users versus controls across three studies. Pooled estimates were obtained using a random-effects model with inverse-variance weighting. Squares represent individual study effects (scaled by weight), horizontal lines indicate 95% CIs, and the diamond represents the pooled effect. Values <1 favor SGLT2i. Moderate between-study heterogeneity was observed ($I^2 = 63.8\%$; $\tau^2 = 0.0166$; $p = 0.0633$).

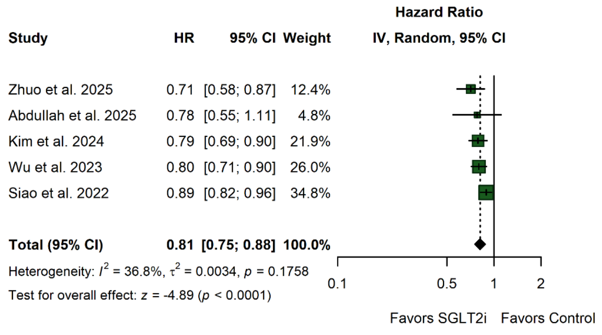


**Figure S4.** Risk of dementia in SGLT2i users. Forest plot of hazard ratios (HRs) with 95% confidence intervals (CIs) comparing SGLT2i users versus controls across five studies. Pooled estimates were obtained using a random-effects model with inverse-variance weighting. Squares represent individual study effects (scaled by weight), horizontal lines indicate 95% CIs, and the diamond represents the pooled effect. Values <1 favor SGLT2i. Low-to-moderate between-study heterogeneity was observed ($I^2 = 36.8\%$; $\tau^2 = 0.0034$; $p = 0.1758$).

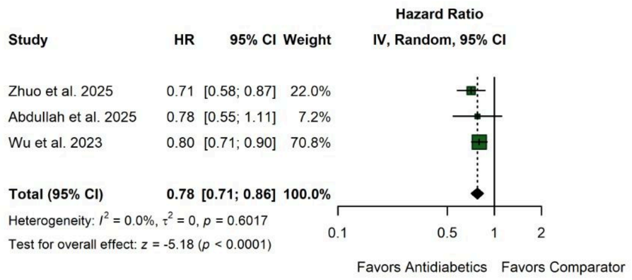


**Figure S5.** Direct comparison between SGLT2i and DPP4i for dementia risk. Forest plot of hazard ratios (HRs) with 95% confidence intervals (CIs) comparing SGLT2i versus DPP4i across three studies. Pooled estimates were obtained using a random-effects model with inverse-variance weighting. Squares represent individual study effects (scaled by weight), horizontal lines indicate 95% CIs, and the diamond represents the pooled effect. Values <1 favor SGLT2i. No between-study heterogeneity was observed ($I^2 = 0.0\%$; $\tau^2 = 0$; $p = 0.6017$).

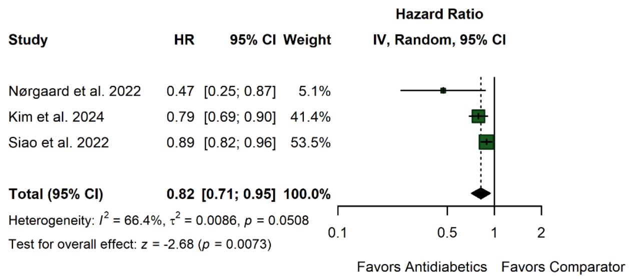


**Figure S6.** Risk of dementia associated with antidiabetic therapy in low-risk-of-bias studies. Forest plot of hazard ratios (HRs) with 95% confidence intervals (CIs) comparing GLP-1RAs and/or SGLT2i versus controls across three studies. Pooled estimates were obtained using a random-effects model with inverse-variance weighting. Squares represent individual study effects (scaled by weight), horizontal lines indicate 95% CIs, and the diamond represents the pooled effect. Values <1 favor antidiabetic therapy. Moderate between-study heterogeneity was observed ($I^2 = 66.4\%$; $\tau^2 = 0.0086$; $p = 0.0508$). The overall effect was statistically significant (HR 0.82, 95% CI 0.71–0.95; $p = 0.0073$).

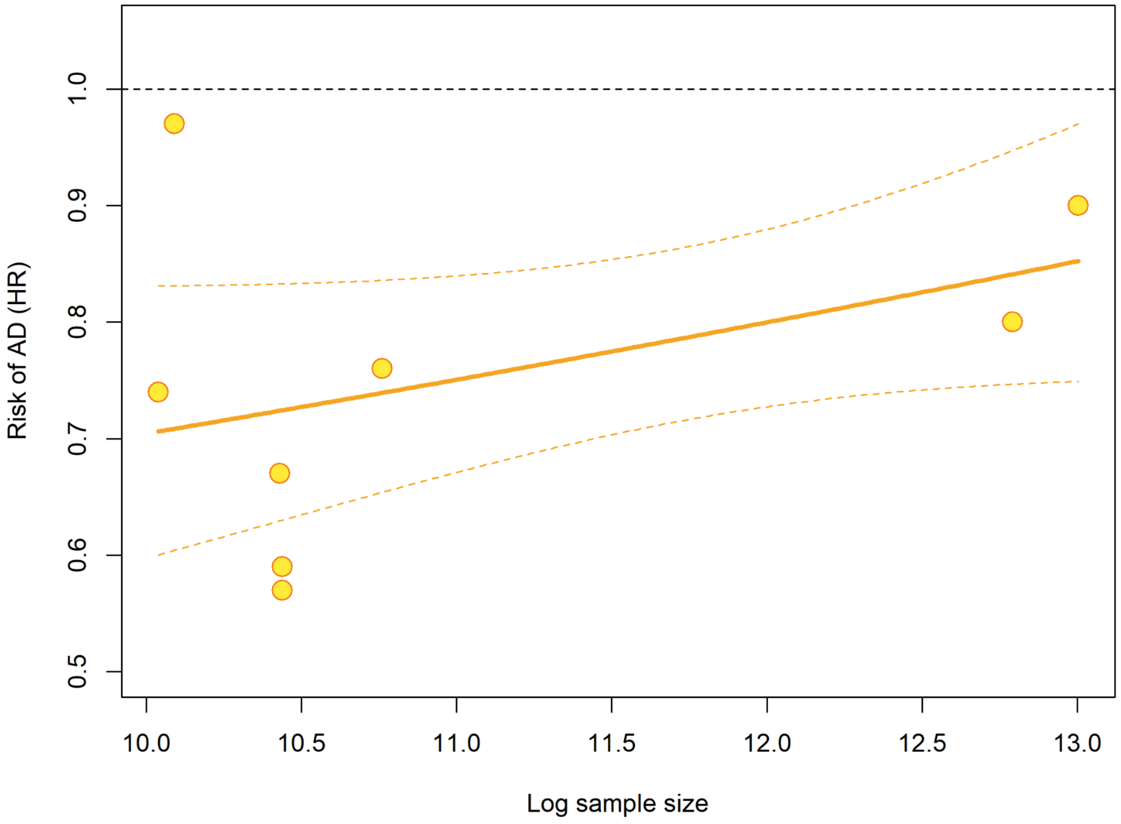


**Figure S7.** Meta-regression analysis evaluating the association between study sample size (log-transformed) and risk of Alzheimer's disease [AD] (expressed as hazard ratios [HRs]). A non-significant positive association was observed ($\beta = 0.064$; SE = 0.038; 95% CI −0.011 to 0.138; $p = 0.096$), with moderate residual heterogeneity ($I^2 = 64.1\%$; $\tau^2 = 0.0073$). Approximately 50.6% of between-study heterogeneity was explained by sample size. These findings suggest a potential small-study effect, although the association did not reach statistical significance.

*HR, hazard ratio; CI, confidence interval; SE, standard error; $\tau^2$, tau-squared; $I^2$, I-squared.*

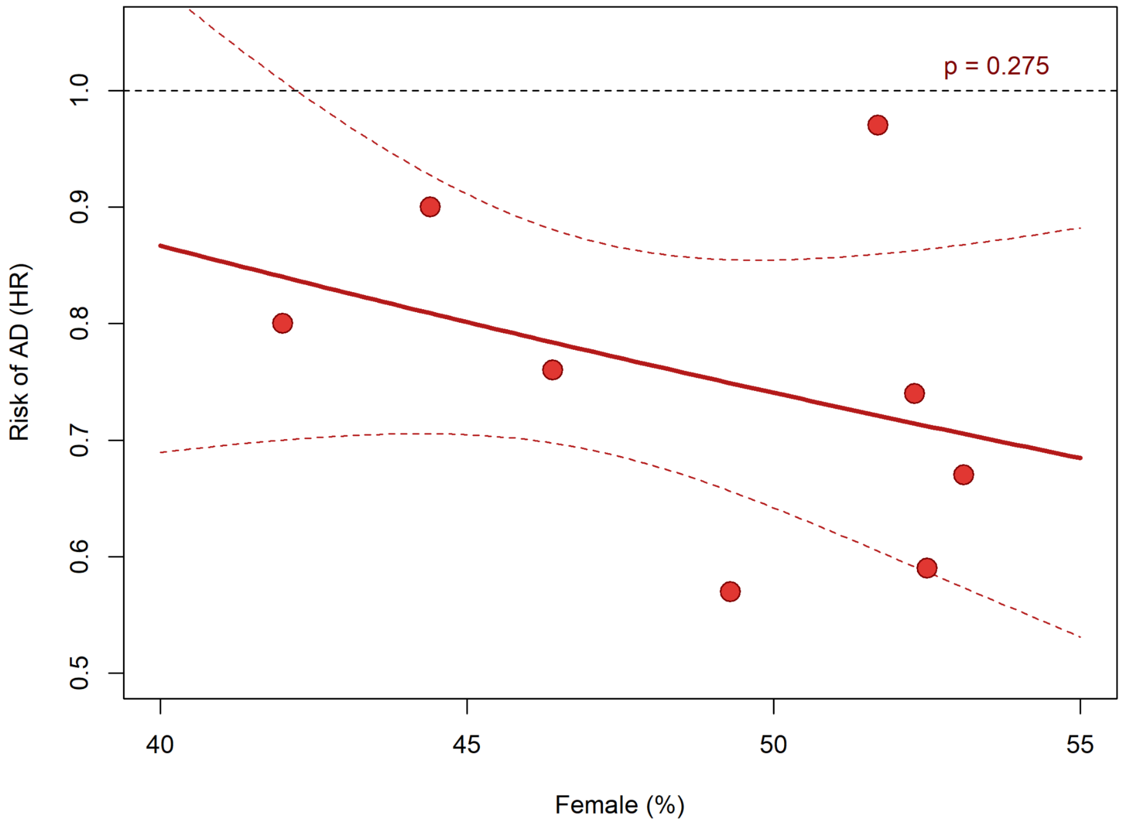


**Figure S8.** Meta-regression analysis evaluating the association between female proportion (%) and risk of Alzheimer's disease [AD] (expressed as hazard ratios [HRs]). No significant association was observed (β = −0.016; SE = 0.014; 95% CI −0.044 to 0.013; p = 0.275), with substantial residual heterogeneity ($I^2$ = 75.9%; $\tau^2$ = 0.014). These findings indicate no evidence that sex distribution influences effect estimates across studies.

*HR, hazard ratio; CI, confidence interval; SE, standard error; $\tau^2$, tau-squared; $I^2$, I-squared.*

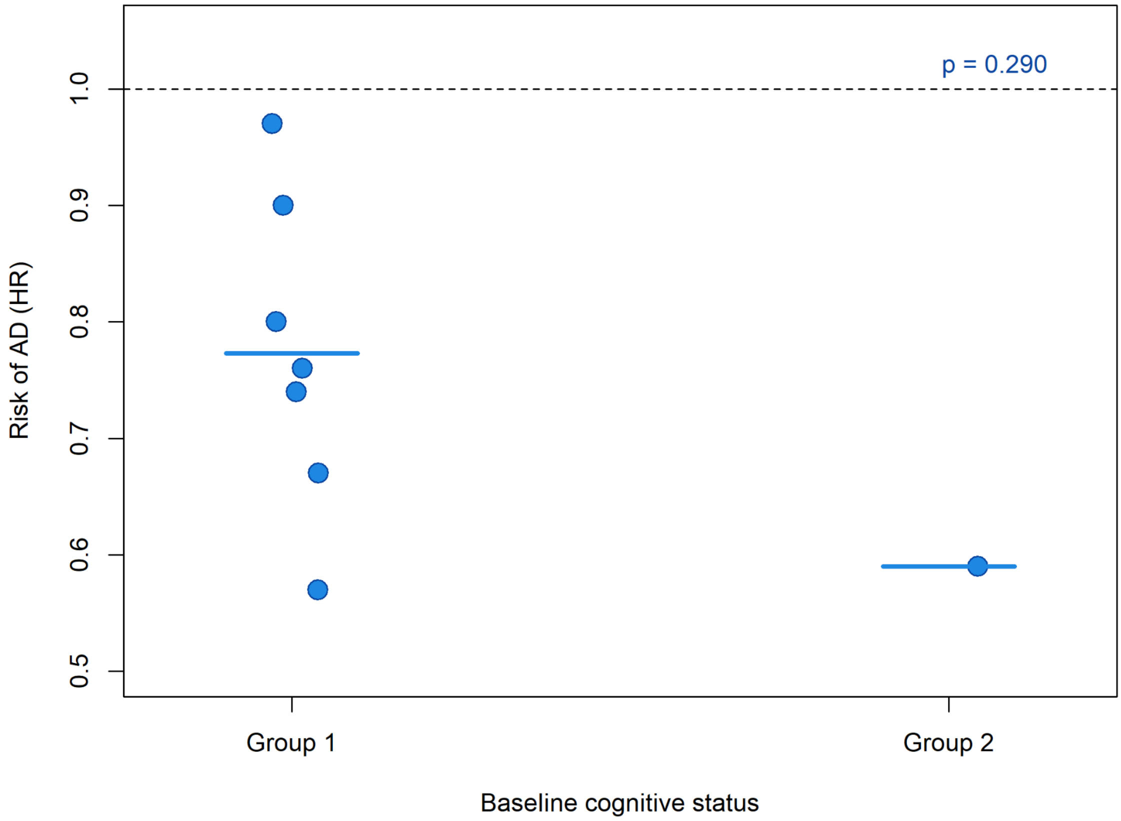


**Figure S9.** Meta-regression analysis evaluating the association between baseline cognitive status and risk of Alzheimer's disease [AD] (HR). No significant association was observed ($\beta = -0.29$; SE = 0.27; 95% CI −0.83 to 0.25; $p = 0.290$), with substantial residual heterogeneity ($I^2 \approx 78.8\%$; $\tau^2 \approx 0.013$). The moderator explained a small proportion of between-study variance ($R^2 \approx 9.8\%$).

*HR, hazard ratio; CI, confidence interval; SE, standard error; $\tau^2$, tau-squared; $I^2$, I-squared.*

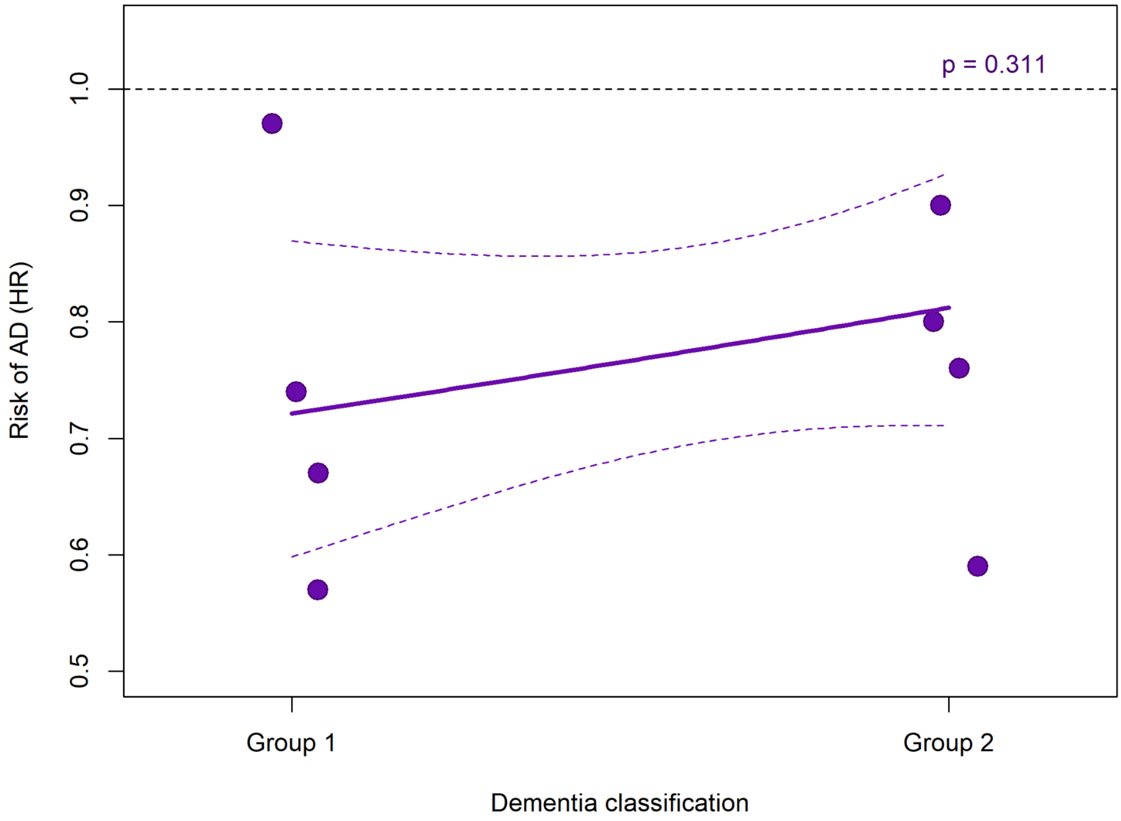


**Figure S10.** Meta-regression analysis evaluating the association between dementia classification and risk of Alzheimer's disease [AD] (HR). No significant association was observed ($\beta = 0.12$; SE = 0.12; 95% CI −0.11 to 0.35; $p = 0.311$), with substantial residual heterogeneity ($I^2 \approx 75.2\%$; $\tau^2 \approx 0.012$). The moderator accounted for a modest proportion of between-study variance ($R^2 \approx 21.9\%$).

*HR, hazard ratio; CI, confidence interval; SE, standard error; $\tau^2$, tau-squared; $I^2$, I-squared.*

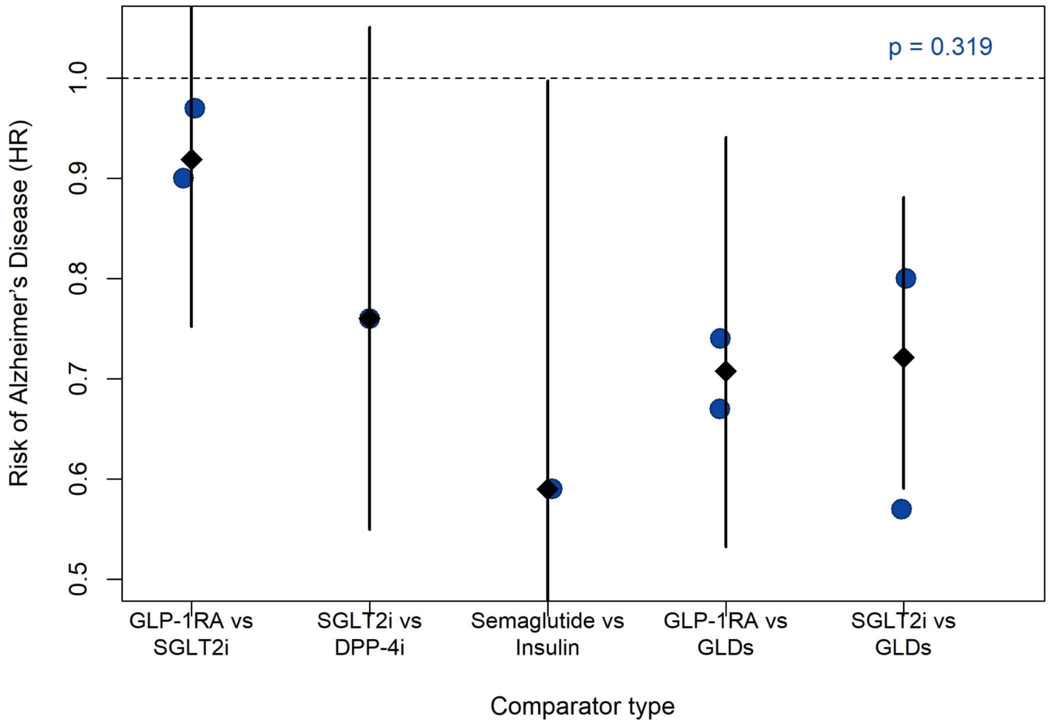


**Figure S11.** Meta-regression analysis evaluating the association between the type of antidiabetic agents and risk of Alzheimer's Disease [AD]. Comparator categories were coded according to predefined groupings across studies. No significant association was observed between comparator type and effect estimates (QM = 4.70; p = 0.319). Residual heterogeneity was moderate ($I^2 \approx 49.5\%$; $\tau^2 \approx 0.014$), with a small proportion of between-study variance explained by the model ($R^2 \approx 6.3\%$). Squares represent individual study estimates, horizontal lines indicate 95% confidence intervals (CIs), and diamonds represent pooled estimates within each comparator category.

*HR, hazard ratio; CI, confidence interval; $\tau^2$, tau-squared; $I^2$, I-squared.*

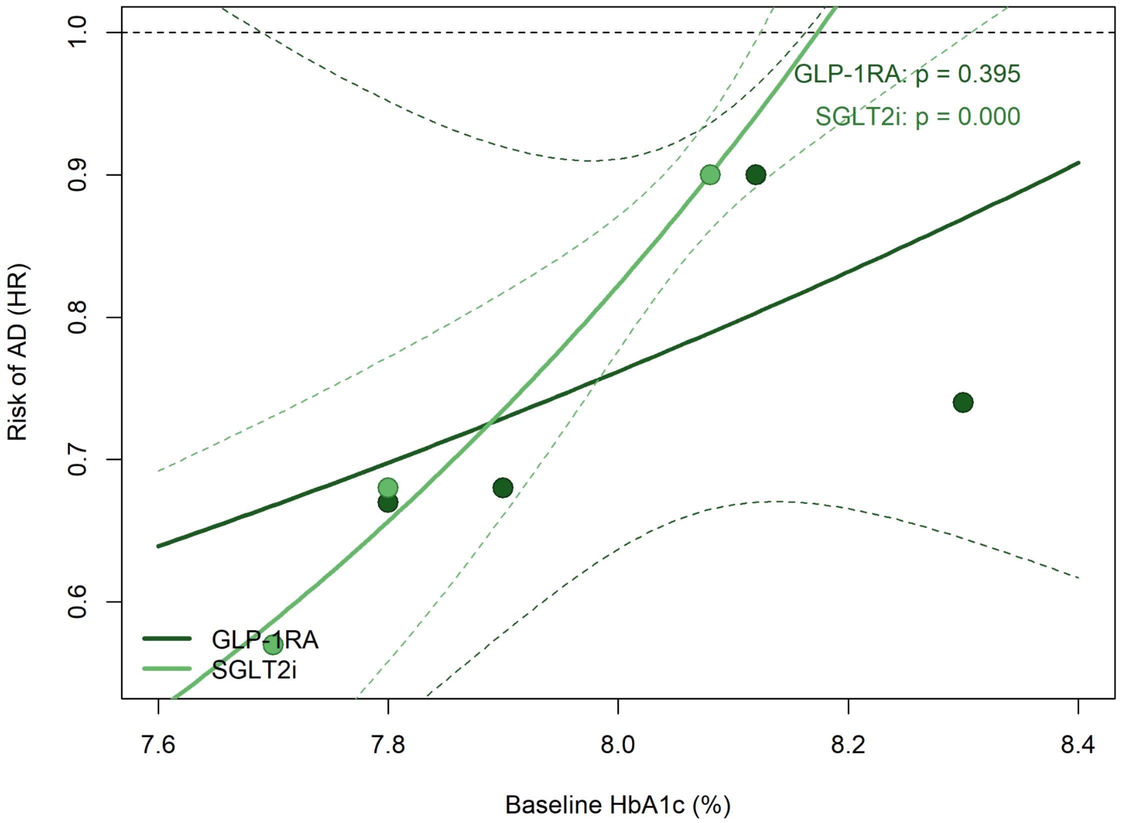


**Figure S12.** Meta-regression analyses evaluating the association between baseline HbA1c (%) and risk of Alzheimer's Disease [AD] (expressed as hazard ratios [HRs]). In the GLP-1RA model, no significant association was observed (β = 0.44; SE = 0.52; 95% CI −0.57 to 1.45; p = 0.395), with moderate residual heterogeneity ($I^2$ = 58.6%; $\tau^2$ = 0.016). In contrast, the SGLT2i model showed a significant positive association (β = 1.13; SE = 0.30; 95% CI 0.53 to 1.72; $p < 0.001$), with no residual heterogeneity ($I^2$ = 0%; $\tau^2$ = 0). These findings indicate no association between HbA1c and effect estimates for GLP-1RA, while the apparent association in the SGLT2i model should be interpreted cautiously, given the small number of studies.

*HR, hazard ratio; CI, confidence interval; SE, standard error; $\tau^2$, tau-squared; $I^2$, I-squared.*

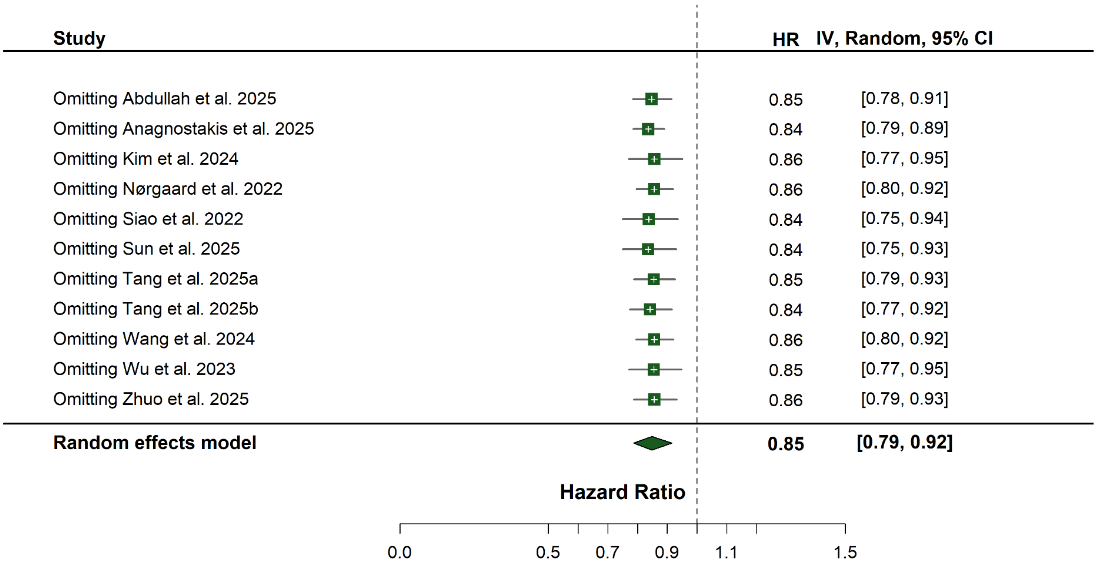


**Figure S13.** Leave-one-out sensitivity analysis for the association between antidiabetic treatment and AD risk.

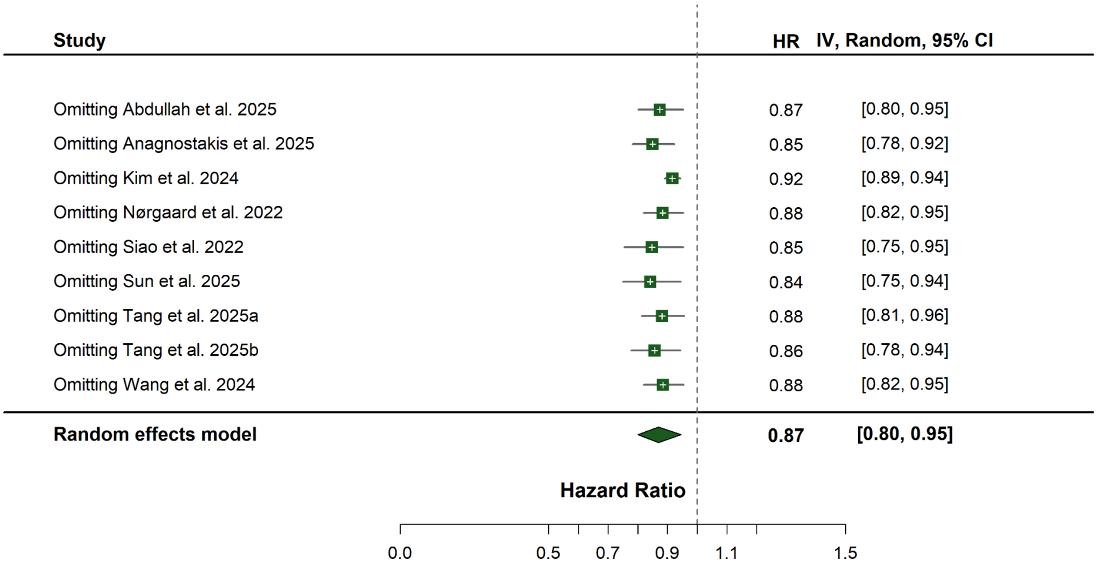


**Figure S14.** Leave-one-out sensitivity analysis for the association between antidiabetic treatment and dementia risk.

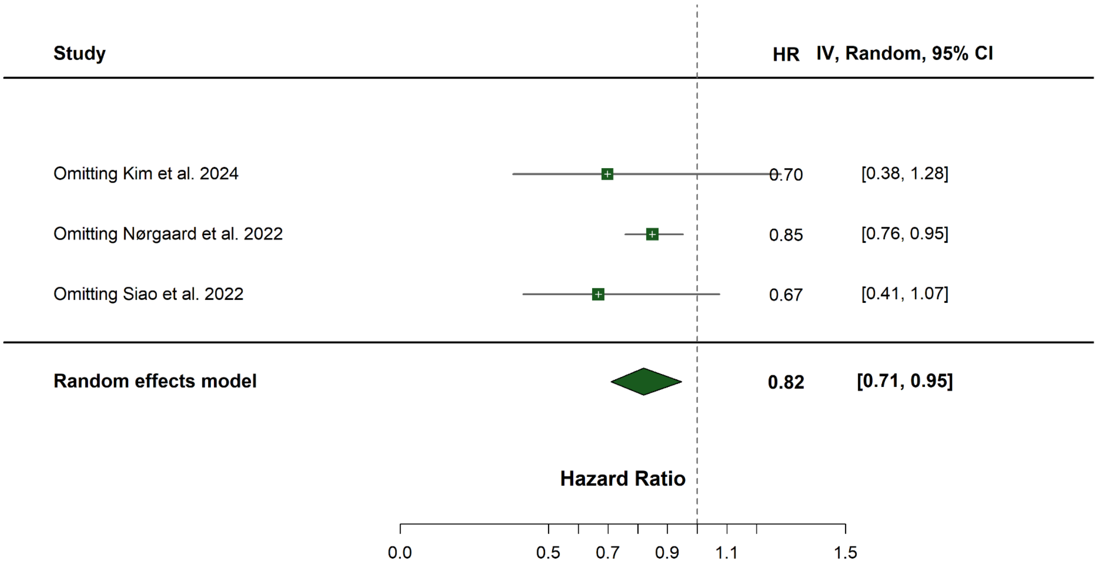


**Figure S15.** Leave-one-out sensitivity analysis for the association between antidiabetic treatment and dementia risk in low-risk-of-bias studies.

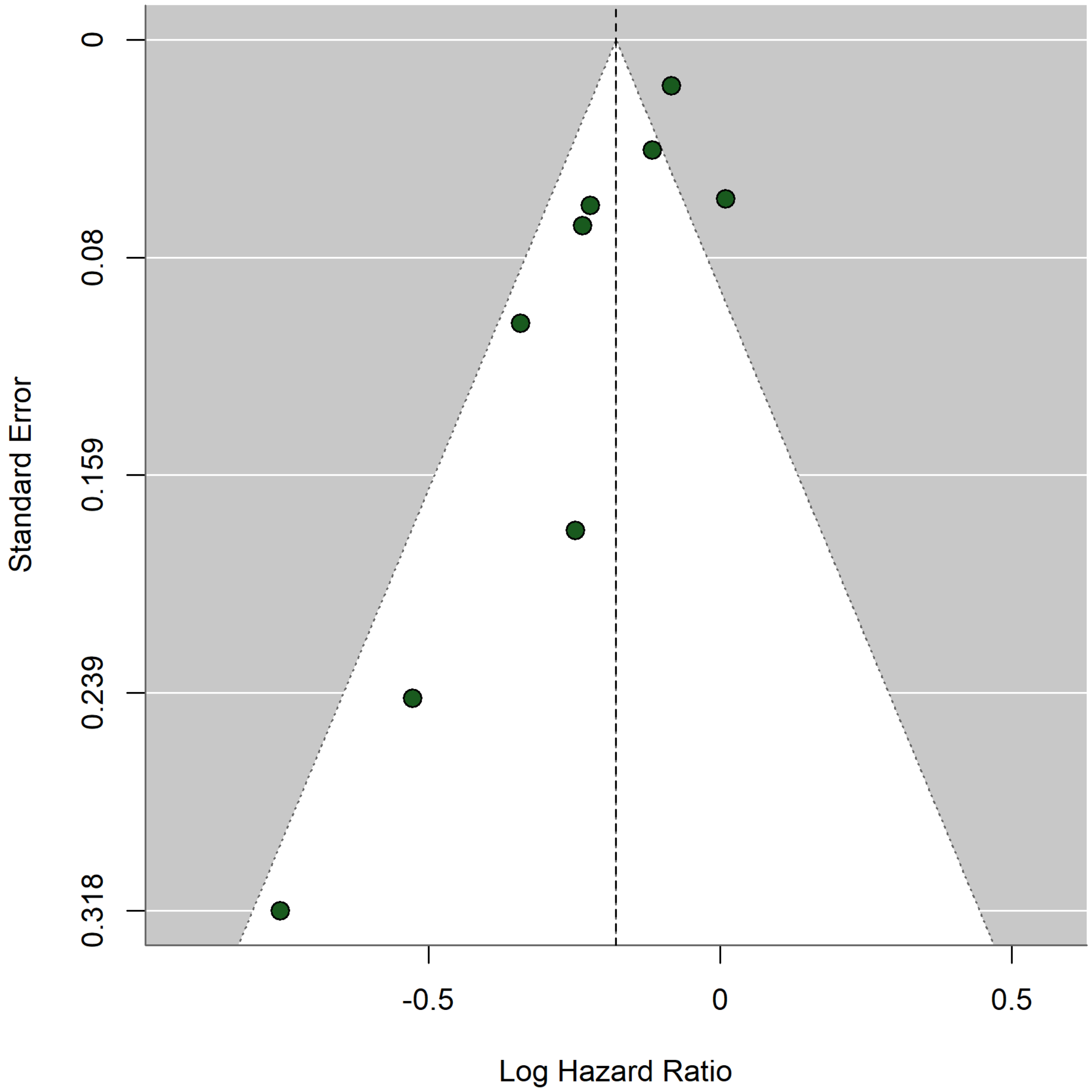


**Figure S16.** Funnel plot of studies reporting hazard ratios (HRs) for Alzheimer's Disease (AD) risk. The plot displays log-transformed HRs against their standard errors. The vertical dashed line represents the pooled effect estimate from the random-effects model (REML), and the shaded triangular region indicates the expected 95% confidence limits in the absence of small-study effects. Visual inspection did not suggest marked asymmetry. Formal statistical testing for funnel plot asymmetry (e.g., Egger's test) was not performed due to the limited number of included studies ($k < 10$).

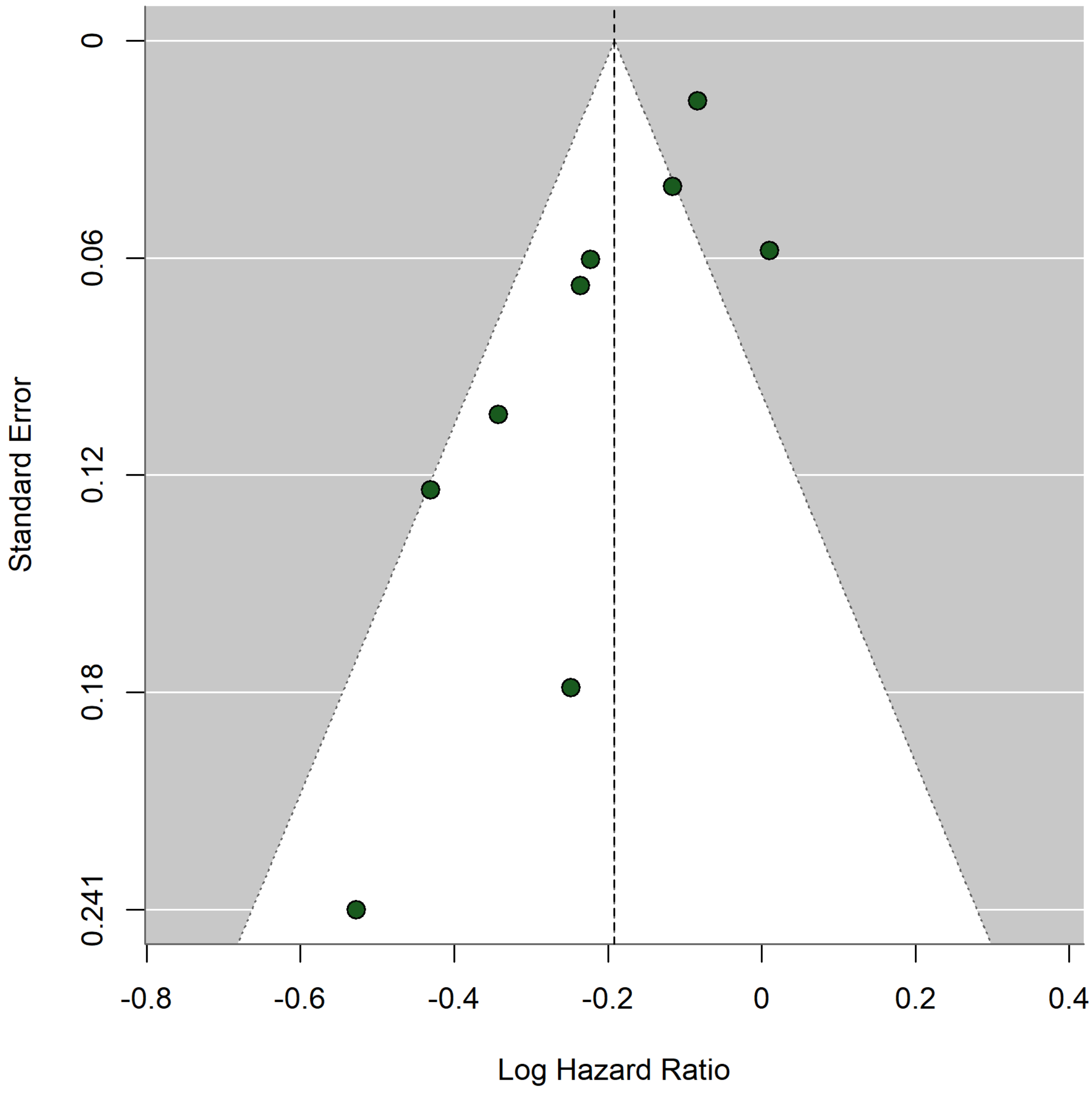


**Figure S17.** Funnel plot of studies reporting hazard ratios (HRs) for dementia risk. The plot displays log-transformed HRs against their standard errors. The vertical dashed line represents the pooled effect estimate from the random-effects model (REML), and the shaded triangular region indicates the expected 95% confidence limits in the absence of small-study effects. Visual inspection did not suggest marked asymmetry. Formal statistical testing for funnel plot asymmetry (e.g., Egger's test) was not performed due to the limited number of included studies ($k < 10$).

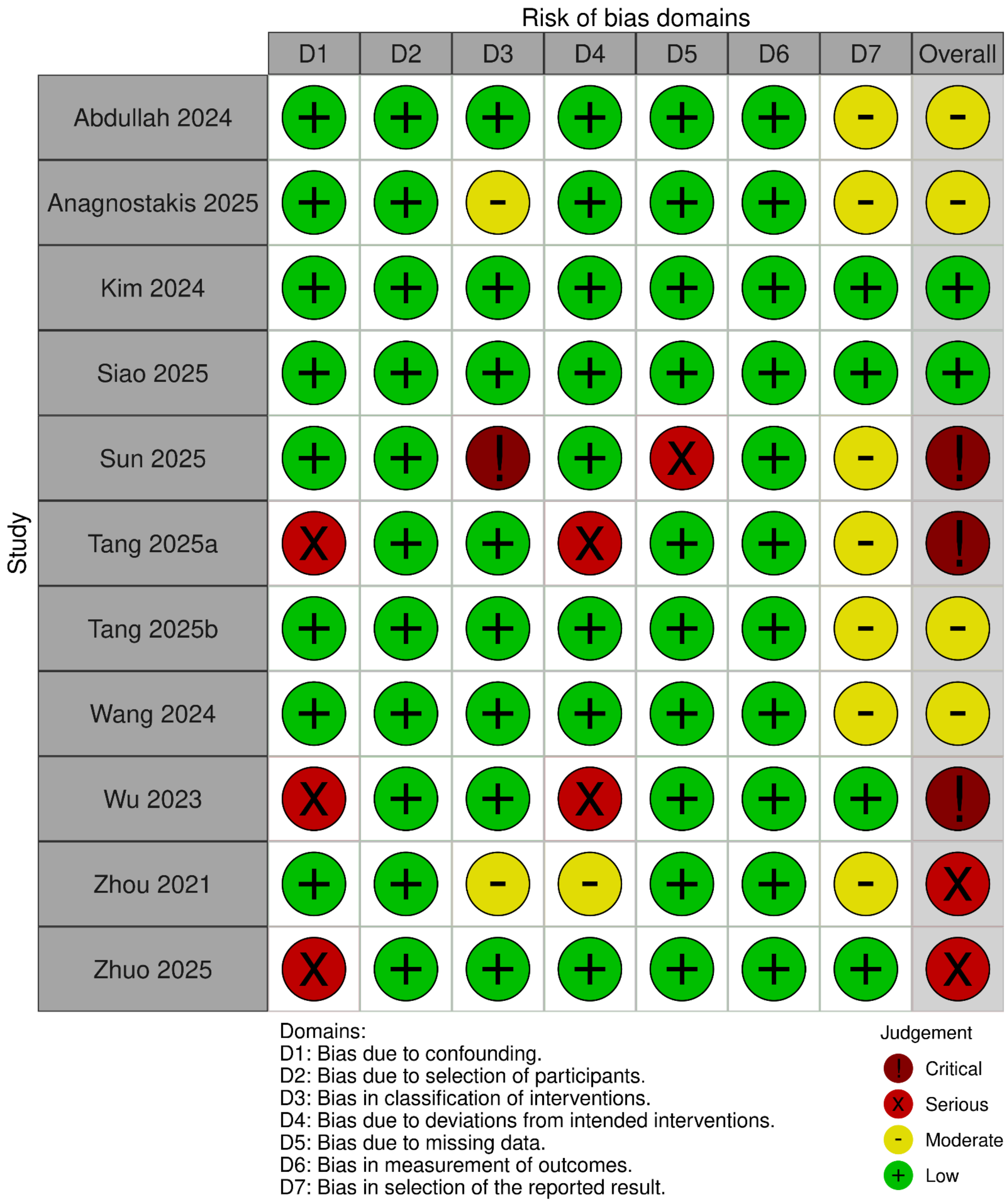


**Figure S18.** ROBINS-I of all included cohorts.

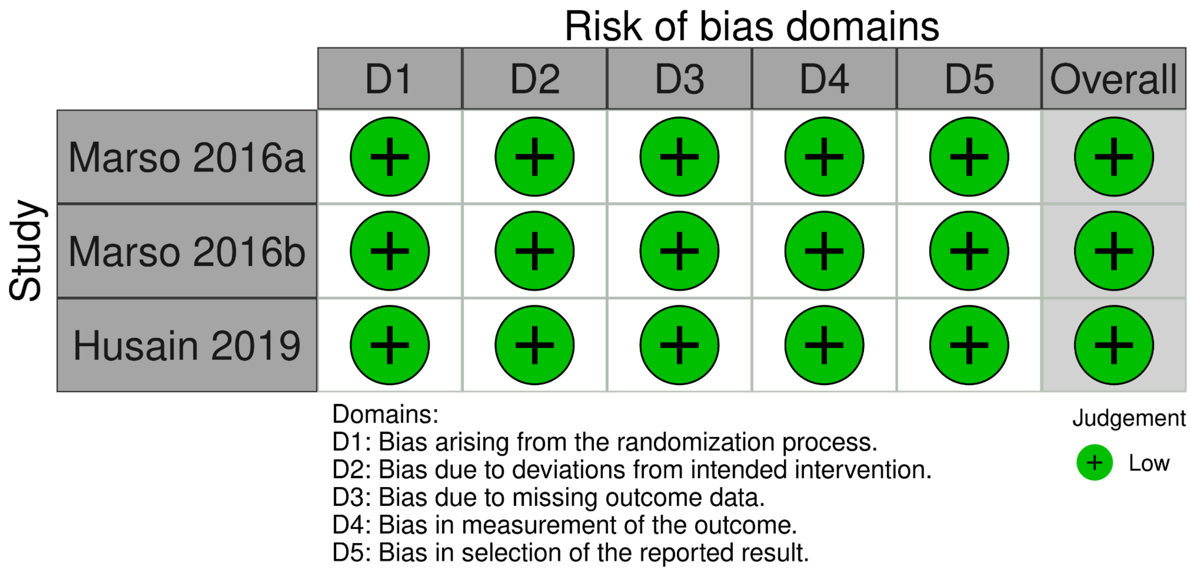


**Figure S19**: RoB 2 tool of the 3 randomized controlled trials (RCTs) pooled in Nørgaard 2022.
